# Investigating causality in the association between DNA methylation and prevalent T2D using a bidirectional two-sample Mendelian Randomization

**DOI:** 10.1101/2022.07.20.22277870

**Authors:** Diana L. Juvinao-Quintero, Gemma C. Sharp, Eleanor Sanderson, Caroline L. Relton, Hannah R. Elliott

**Author notes:** Corresponding author: Hannah R Elliott. Co-Corresponding author: Diana L. Juvinao-Quintero.

## Abstract

**Aim:** Several studies have identified associations between type 2 diabetes (T2D) and DNA methylation (DNAm). However, the causal role of these associations remains unclear. This study aims to provide evidence for a causal relationship between DNA methylation and T2D.

**Methods:** We implemented a bidirectional two-sample Mendelian randomization (2SMR) to evaluate causality at 58 CpG sites previously detected in a meta-analysis of epigenome-wide association studies (meta-EWAS) of prevalent T2D in Europeans. We retrieved genetic proxies for T2D and DNAm from the largest GWAS available. We also used data from the Avon Longitudinal Study of Parents and Children (ALSPAC, UK) when associations of interest were not available in the larger datasets. We identified 62 independent SNPs as proxies for T2D, and 39 methylation quantitative trait loci or mQTL as proxies for 30 of the 58 T2D-related CpGs. We applied correction for multiple testing using Bonferroni and inferred causality based on a *P* < 1.0×10^−3^ or *P* < 2.0×10^−3^ for the T2D⟶ DNAm direction, and the opposing DNAm ⟶ T2D direction of the 2SMR, respectively.

**Results:** We found strong evidence of causality of DNAm at cg25536676 (*DHCR24*) on T2D, where an increase in transformed residuals of DNAm at this site were associated with 43% (OR=1.43, 95%CI=1.15-1.78, *P*=0.001) higher risk of T2D. We infer a likely causal direction for the remaining CpG sites assessed. *In silico* analyses showed that CpGs analyzed were enriched for eQTMs, and for specific traits dependent on the direction of causality predicted by 2SMR.

**Conclusions:** We identified one CpG mapping to a gene related with the metabolism of lipids (*DHCR24*), as a novel causal biomarker for the risk of T2D. CpGs within the same gene-region have previously been associated with T2D-related traits in observational studies (BMI, waist circumference, HDL-cholesterol, insulin) and in MR analyses (LDL-cholesterol). Thus, we hypothesize that our candidate CpG in *DHCR24* may be a causal mediator of the association between known modifiable risk factors and T2D. Formal causal mediation analysis should be implemented to further validate this assumption.

## INTRODUCTION

In recent years, there has been a growing interest in understanding the role of DNA methylation (DNAm) in the context of type 2 diabetes (T2D). In epidemiological studies, associations have been identified between DNA methylation and both prevalent [1-8] and incident T2D [9-14]. Associations between DNAm and T2D may arise through different mechanisms. For example, changes in DNAm may be causal on T2D. Conversely, T2D may induce consequent changes in DNAm. It is also possible that the observed association between DNAm and T2D arises due to confounding from a third factor that is independently associated with both T2D and DNAm. Defining the causal relationship between DNAm and T2D is important because it provides new information about the molecular pathways involved in disease incidence, and potentially progression, providing new insights into targets for intervention.

Prior studies have investigated the causal direction of association between DNA methylation and T2D using different study designs. In the context of incident T2D, the causal link between 18 incident T2D associated CpG sites and T2D was assessed using Mendelian randomization [11]. mQTLs were available for 16 of the 18 T2D associated CpG sites. There was nominal evidence for a direct causal association of cg00574958 (*CPT1A)* on T2D [11]. The authors also assessed the causal effects of BMI and glycaemic traits on methylation at the 18 CpG sites associated with incident T2D but found no evidence to support this causal pathway [11]. In the context of prevalent T2D, a study of 232 cases and 197 controls from a Korean cohort identified 12 CpG sites associated with T2D and measured the association between these CpG sites and metabolic traits in a further 1018 individuals [7]. MR analyses revealed that there was a likely causal effect of fasting glucose on cg00574958 (*CPT1A)*. There was no evidence to suggest causality of T2D on methylation in this study [7]. GrimAge, an estimate of epigenetic ageing derived from DNA methylation, has been shown to mediate the relationship between cumulative obesity and T2D [13].

Using the genotype as causal anchors to assess future risk of T2D, DNA methylation did not appear to be on the causal pathway between known T2D genetic risk variants and T2D, with the exception of *KCNQ1* [15]. A further study identifying T2D genetic risk variants which were also *cis-*mQTLs, assessed the causal pathway from methylation to T2D using MR. This study identified CpG sites at four loci (*HNF1B, KCNJ11, IGF2BP2 and WFS1*) that were likely causally associated with future risk of T2D [16].

While efforts have been made to determine the causal direction of associations between DNA methylation and T2D, conclusions remain sparse and inconsistent. In part, this is because previous studies have used modestly sized datasets (i.e., <1000 individuals). The first aim of this study was to comprehensively investigate the causal direction of associations between DNAm and prevalent T2D. This analysis was conducted using a bi-directional Mendelian randomization approach in CpG sites previously identified in a meta-EWAS of prevalent T2D. This meta-EWAS is the largest meta-EWAS of prevalent T2D currently available, including 340 T2D cases and 3,088 controls from four European cohorts. The second aim of this study was to determine the functional role of CpGs stratified by their likely causal direction of effect on T2D. We hypothesized that the functional role of CpGs would be different dependent on their causal direction of effect, potentially highlighting key molecular pathways for interventions.

## METHODS

### Study samples

#### Forward two-sample MRT2D as causal of differences in DNAm at candidate CpGs

We extracted 148 genetic variants (SNPs) associated with T2D from four GWAS in the DIAGRAM consortium [17] (Supplementary Table 1), three of them were multi-ethnic [18-20] and one was European [21]. We selected a SNP if it was (1) identified as an index variant in a GWAS meta-analysis, (2) or a SNP found within a 99% credible set from an index variant, and with equal or higher posterior probability than the index variant, or a (3) SNP identified in the exome mapping close to a well-established locus for T2D, with minor allele frequency (MAF) > 0.05. Two levels of significance were considered in the selection of SNPs: a genome-wide significance threshold *P* < 5.0×10^−8^, and a locus-wide significance *P* < 1.0×10^−5^ (GCTA joint regression model). SNPs were excluded if they had incomplete data for the risk allele, the reported effect estimate (OR), the P-value, or if MAF < 0.05. After quality control in MR-Base [22], the initial list of 148 T2D SNPs was reduced to 62 not highly correlated SNPs (LD < 0.2) [23] (Supplementary Table 2) with high-quality genotype data in our outcome sample (i.e., ALSPAC-ARIES).

We used data from ALSPAC-ARIES [24-26] (Supplementary Material S1) to extract estimates of the association between 62 T2D SNPs and DNAm levels at 58 CpG sites previously associated with prevalent T2D [6]. For this analysis, we included cross-sectional data from 1,243 middle-aged participants (mean age 49.1 years, range 31 to 75 years) irrespective of their T2D status. We could not use GoDMC [27] as our outcome sample because T2D SNPs, or related SNPs in high LD (r > 0.6), were associated with CpGs of interest at *P* > 10^−8^, which was the maximum P-threshold used by GoDMC to report summary data. Figure 1 shows the study design used to conduct the forward two-sample MR analysis.

**Figure 1.**
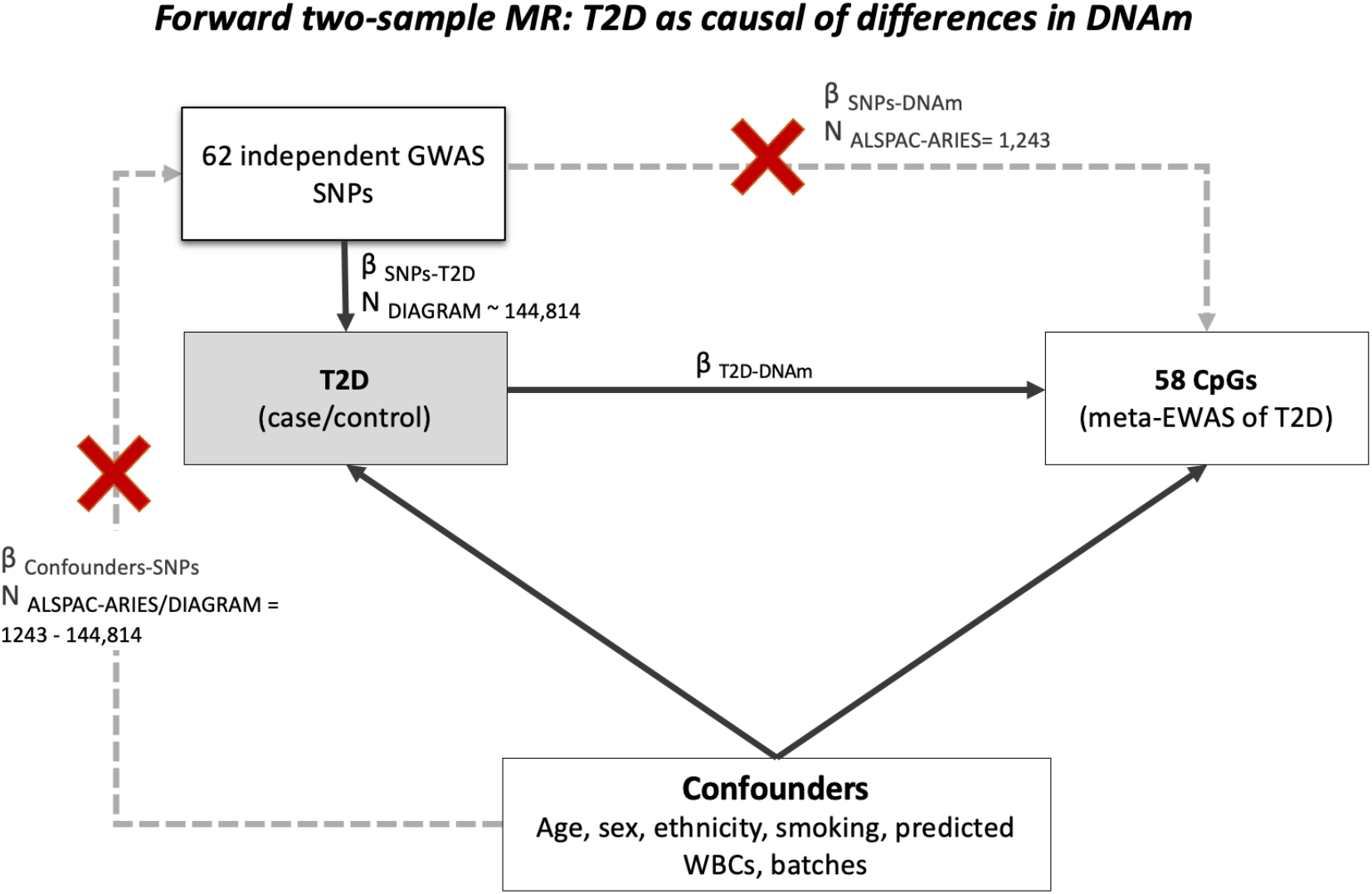
Study design of the forward two-sample MR (2SMR) investigating the causal effect of T2D on differences in DNAm at 58 CpG sites. CpGs were previously identified in association with T2D in a meta-analysis of EWAS (meta-EWAS). IV: instrumental variable, WBC: white-blood cells.

#### Reverse two-sample MR: DNAm at candidate CpGs as causal risk factors for T2D

Using summary data from GoDMC [28], we retrieved 41 mQTL associated with DNAm in blood at 31 of the 58 CpG sites previously reported in a meta-EWAS of T2D (Supplementary Material S5). We selected mQTL with a *P* < 10^−8^ for *cis*- (SNP < 1 Mb from CpG site) and a *P* < 10^−14^ for *trans*-mQTL (SNP > 1 Mb from CpG site). In total, 5 of the 41 mQTL identified were *trans*-mQTL. When there was multiple mQTL for a single CpG, we used *clumping* to select only those that were independent (Supplementary Material S6). Data extracted for each mQTL was obtained using a fixed-effect meta-analysis, with effect estimates interpreted as a unit change in inverse-normal transformed residuals of DNA methylation, per additional effect allele.

We verified that mQTL SNPs used in the reverse MR were independent of SNPs for T2D in the forward MR by using a LD threshold r^2^ < 0.01. The correlation between T2D SNPs and mQTL was calculated using Ldlink (https://ldlink.nci.nih.gov/) [29], selecting as the reference panel genetic data from European samples in the 1,000 Genomes project phase 3. The explanation for selecting independent instruments across the two directions of the bidirectional MR, is that this measure reduces the chances of bias in the causal estimate (i.e., pleiotropic effects or reverse causation) [30].

As our outcome sample, we selected the two largest T2D GWAS available at the time [18, 31], one corresponding to a trans-ethnic meta-GWAS published by Mahajan *et al*. 2014 [18], and the second a GWAS published by Wood *et al*. 2016 [31] using data from the UK Biobank. Both datasets for the outcome were available in MR-Base [22]. Associations of mQTL with T2D were extracted using the MRInstruments and TwoSampleMR R packages. When information for a mQTL was not available in the outcome sample, MR-Base looked for alternative SNPs with a LD > 0.8 with the target mQTL SNP. After data harmonization in MR-Base, we were able to successfully extract outcome data for 39 of the initial 41 mQTL SNPs typing 30 meta-EWAS CpGs. We present the study design used to conduct the reverse two-sample MR in Figure 2.

**Figure 2.**
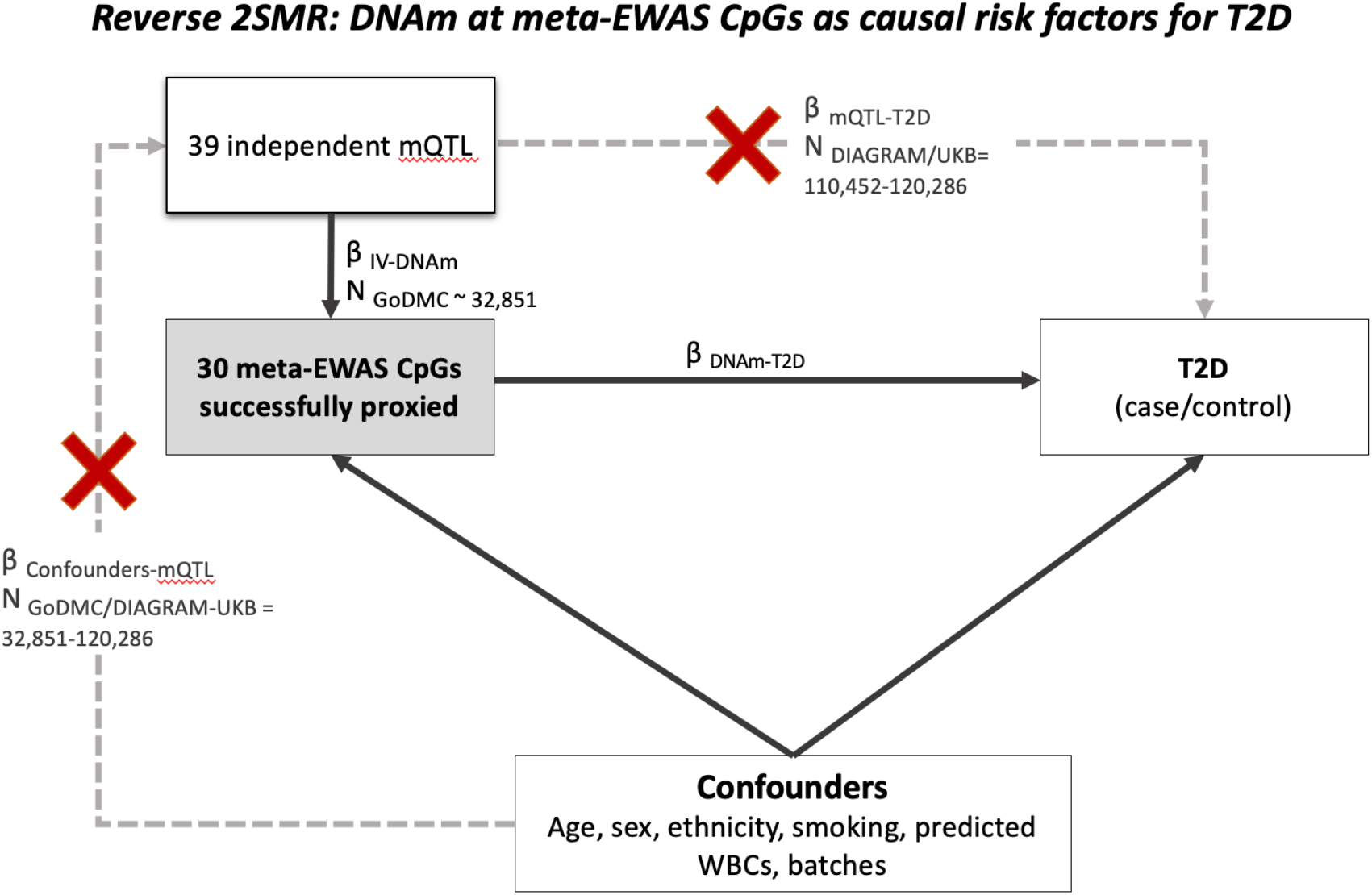
Study design of the reverse two-sample MR (2SMR) investigating the causal effect of differences in DNAm on T2D risk. 58 CpGs were previously identified in association with T2D in a meta-analysis of EWAS (meta-EWAS), but only 30 of them were proxied by a mQTL SNP in GoDMC with available GWAS data for T2D in MR-Base. IV: instrumental variable, PCs: genetic principal components, UKB: UK-Biobank.

**Figure 3.**
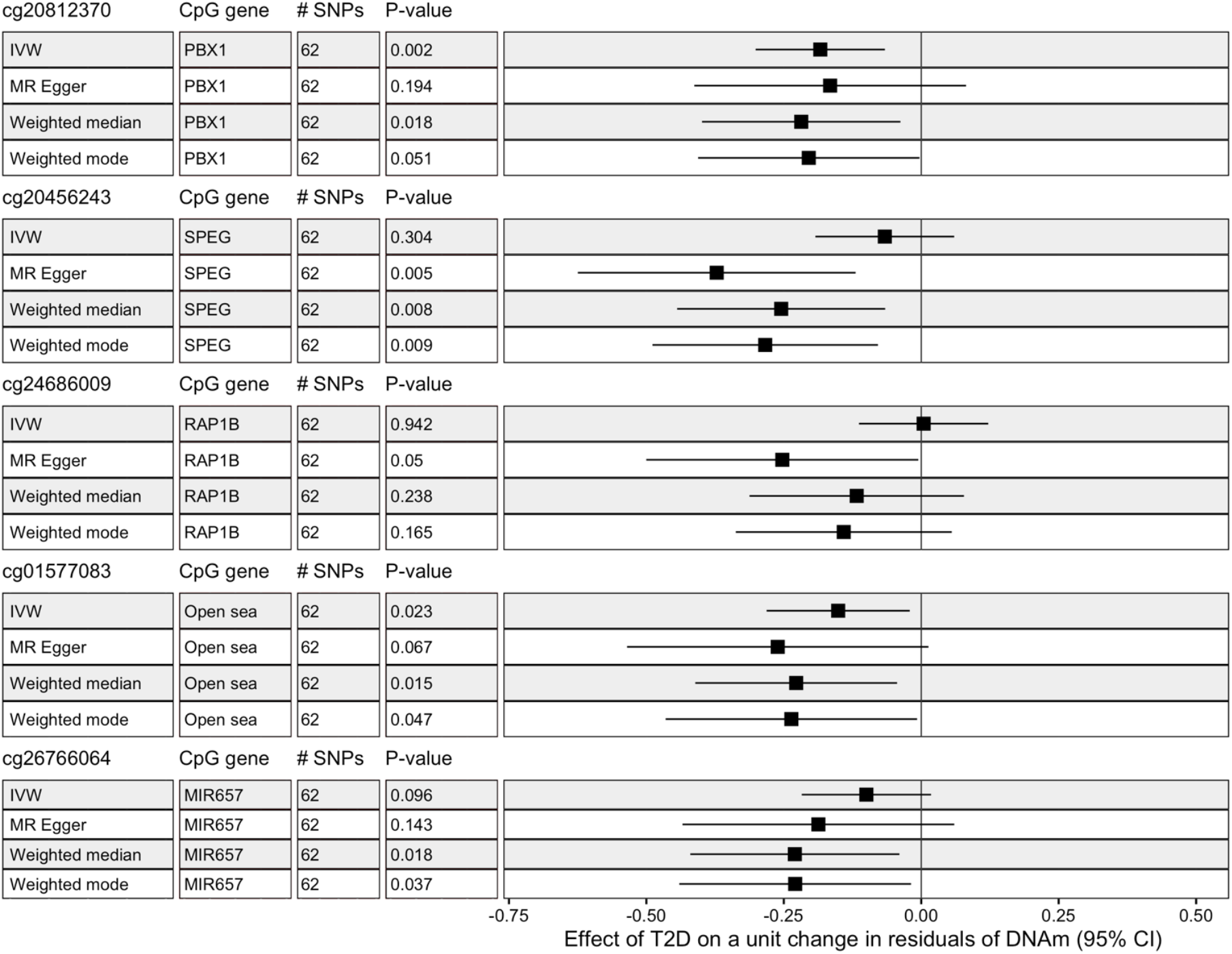
Forest plot showing causal effect estimates from a two-sample MR looking at the effect of prevalent T2D on the difference in DNAm at five meta-EWAS CpGs identified with the strongest evidence of causality in the forward 2SMR. Mean causal effect for each CpG and MR method is represented by the black square, and the line across is the 95% confidence interval. IVW: inverse-variance weighted random effect regression. # SNPs: number of valid single nucleotide polymorphisms used as proxies for T2D in the forward MR analysis. P-value: unadjusted P-value. Associations were borderline significant at unadjusted-P < 0.05, or significant at adjusted-P < 0.001.

### Statistical analyses

#### Forward two-sample MR: Conducting a SNP-CpG analysis in ALSPAC-ARIES

We measured the association between 62 independent T2D SNPs and 58 CpG sites previously reported in a meta-EWAS of T2D using a standardized the protocol [27]. We conducted linear regressions with the genotype for T2D as the exposure and DNAm at each CpG site as the outcome using an additive genetic model. Briefly, in the first stage of the analysis, we extracted complete genetic and DNAm data for 1,243 middle-adults in ALSPAC-ARIES, and we performed QC on each dataset separately (Supplementary Material S2-S4). For genome-wide SNPs and CpG sites that surpassed QC, we selected only those corresponding to T2D SNPs and meta-EWAS CpGs, respectively, to conduct regression analyses using the MatrixeQTL R package. We tested the direct effect of the genotype for T2D on DNAm at meta-EWAS CpGs at *P* < 1.4×10^−5^ or α=0.05/62 T2D SNP*58 CpGs. We interpreted effect estimates of the SNP-CpG analysis as the difference in residuals of DNAm (inverse-normal transformed values), per additional risk allele for T2D.

#### Mendelian randomization and MR-Base analysis

Mendelian randomization is a statistical method used to infer causality in observational associations. Detailed description of the method has been documented elsewhere [32-34]. Briefly, MR uses the genotype as an unconfounded “causal anchor” to estimate causal effects of the exposure on the outcome unbiased from the effects of confounders [32]. For this study, we used a two-sample MR, where summary data for the association of the genetic instrument(s) with the exposure and the genetic instrument(s) with outcome, was retrieved from two independent but comparable populations [35], both with moderate power. A two-sample MR analysis can be easily automated using MR-Base. This platform serves as a repository of curated GWAS, and as an analytical tool to perform various MR analyses [22]. Using MR-Base, we implemented tools available in the TwoSampleMR (version 0.5.6), MRInstruments (version 0.3.2) and MendelianRandomization (version 0.5.1) R packages to conduct causal analyses [22, 36]. Further detail is provided in the Supplementary Material S6.

### Determining true direction of association

For associations analyzed bidirectionally, we inferred the likely causal direction using the causal effect estimate with the smallest P-value, that was also consistent with the direction of association found in the observational analysis (i.e., meta-EWAS of T2D). “Inconclusive” associations with bidirectional MR data had a P-value > 0.1 in each direction of the analysis. For associations with analyzed MR data in a single direction only, we regarded them as “inconclusive-single direction”.

### Functional Inspection of MR signals using publicly available databases

We classified meta-EWAS CpGs into three subgroups based on their most likely causal direction of association with T2D: 1) T2D causal of DNAm variation, 2) DNAm causal of T2D risk, and 3) inconclusive direction of association. We looked for expression quantitative trait methylation loci in *cis* (*cis*-eQTM) associated with meta-EWAS CpGs in the BIOS QTL browser (https://genenetwork.nl/biosqtlbrowser/) [37]. In addition, results from the EWAS Catalog [38] were grouped into related phenotypes [39] and tested for enrichment among the 58 meta-EWAS CpGs analyzed. Enrichment of each CpG subgroup for specific phenotypes was reported using odd ratios (OR) and 95% confidence intervals (95%CI). P-values of the enrichment analysis were one-sided. We considered evidence of enrichment at *P* < 0.05/traits analyzed.

## RESULTS

### Forward 2SMR: T2D was suggestively associated with lower DNAm at cg20812370 (*PBX1*), previously identified in a meta-EWAS of T2D

Summary statistics of the associations between 62 SNPs and T2D are shown in the Supplementary Table 2, while association estimates between T2D SNPs and 58 CpGs are presented in the Supplementary Table 3.

Implementing a 2SMR, we observed suggestive evidence of causality (adjusted *P* < 0.001 or unadjusted *P* < 0.05) between T2D and lower levels of DNAm at the CpGs cg20812370 (*PBX1*) (*P*=0.002) and cg01577083 (open sea) (*P*=0.023). The causal effects at both CpGs were directionally consistent with results of the observational analysis (meta-EWAS), but absolute effect sizes were always larger in the MR compared to the observational analysis (Table 1). For associations at cg20812370 (*PBX1*) and cg01577083, we had little evidence of heterogeneity in the effect of 62 T2D SNPs on DNAm levels based on results of the Cochran’s Q estimate (Q range 52.9 - 74.8, *P* range 0.10 - 0.75).

**Table 1.** Observational association and estimates of causal effect between prevalent T2D and difference in DNAm at five CpGs observed with the strongest evidence of causality in the forward 2SMR. Observational estimates were obtained from a previous meta-analysis of EWAS of T2D.

| CpG | Chr | Gene | Meta-EWAS of T2D |  |  | MR IVW regression |  |
| --- | --- | --- | --- | --- | --- | --- | --- |
|  |  |  | Effect (95%CI) | P | N | Effect (95%CI) | P |
| cg20812370 | 1 | *PBX1* | -0.007(-0.009, -0.004) | 7.40E-07 | 3428 | -0.184(-0.301, -0.066) | 0.002 |
| cg20456243 | 2 | *SPEG* | -0.007(-0.011, -0.004) | 9.99E-06 | 3428 | -0.066(-0.193, 0.06) | 0.304 |
| cg24686009 | 12 | *RAP1B* | -0.002(-0.003, -0.001) | 1.19E-06 | 3428 | 0.004(-0.113, 0.122) | 0.942 |
| cg01577083 | 16 | *Open sea* | -0.011(-0.016, -0.006) | 7.93E-06 | 3428 | -0.151(-0.281, -0.021) | 0.023 |
| cg26766064 | 17 | *MIR657* | -0.007(-0.009, -0.004) | 5.17E-06 | 3428 | -0.100(-0.217, 0.018) | 0.096 |

Meta-EWAS of T2D: meta-analysis of epigenome-wide association studies of prevalent T2D [6]. IVW: inverse-variance weighted random-effect regression. Associations were considered significant at $P < 0.001$ or $\alpha = 0.05/58$ CpGs analyzed in the forward 2SMR.

When we used the MR-Egger, weighted mode and weighted median as sensitivity analyses, we observed four CpGs with suggestive evidence of causality with T2D at *P* < 0.05, including previously detected cg20812370 (*PBX1*) and cg01577083 (open sea), in addition to cg20456243 (*SPEG*) and cg26766064 (*MIR657*) (Table 2). In all cases, magnitude and direction of the effect estimate was similar across analyses, while smaller P-values were generally seen when using the weighted median compared to the weighted mode and MR-Egger regressions, respectively. Results of these sensitivity analyses were directionally consistent with estimates of the IVW regression, and with the observational analysis (meta-EWAS).

**Table 2.** Estimates of causal effect between prevalent T2D and difference in DNAm at five CpGs using additional MR sensitivity methods for the forward 2SMR.

| CpG | Chr | Gene | MR-Egger regression |  | Weighted Median |  | Weighted Mode |  |
| --- | --- | --- | --- | --- | --- | --- | --- | --- |
|  |  |  | Effect (95%CI) | P | Effect (95%CI) | P | Effect (95%CI) | P |
| cg20812370 | 1 | *PBX1* | -0.166(-0.413, 0.082) | 0.194 | -0.218(-0.399,-0.038) | 0.018 | -0.205(-0.406,-0.003) | 0.051 |
| cg20456243 | 2 | *SPEG* | -0.372(-0.625, -0.119) | 0.005 | -0.255(-0.444,-0.065) | 0.008 | -0.284(-0.489,-0.079) | 0.009 |
| cg24686009 | 12 | *RAP1B* | -0.253(-0.5, -0.005) | 0.050 | -0.117(-0.312,0.078) | 0.238 | -0.141(-0.337,0.056) | 0.165 |
| cg01577083 | 16 | *Open sea* | -0.261(-0.535, 0.013) | 0.067 | -0.227(-0.411,-0.044) | 0.015 | -0.236(-0.465,-0.007) | 0.047 |
| cg26766064 | 17 | *MIR657* | -0.187(-0.434, 0.06) | 0.143 | -0.230(-0.420,-0.040) | 0.018 | -0.229(-0.44,-0.019) | 0.037 |

MR-Egger: sensitivity analysis to account for horizontal pleiotropy. Weighted median and weighted mode: sensitivity analyses that allow for some instruments to be invalid, while generating an unbiased causal estimate with the set of valid proxies. Associations were considered significant at $P < 0.001$ or $\alpha = 0.05/58$ CpGs analyzed in the forward 2SMR.

Overall, in the forward the 2SMR, we found no evidence of weak instrument bias based on values of the F-statistic ranging from 19.7 to 274.8 (strong instrument if F-statistic > 10).

### Reverse 2SMR: Elevated DNAm at the CpG cg25536676 in *DHCR24* was associated with increased risk of prevalent T2D

Summary estimates of the association between 39 mQTL SNPs and 30 meta-EWAS CpGs are shown in the Supplementary Table 4. Association estimates of 39 mQTL SNPs with T2D are presented in the Supplementary Table 5. Overall, none of these mQTL were directly associated with T2D with GWAS significance (*P* < 5.0×10^−8^). Four mQTL tagging the CpGs cg08857797 (*VPS25*), cg00144180 (*HDAC4*), cg16765088 and cg25536676 (*DHCR24*), were suggestively associated with T2D with unadjusted GWAS *P* < 0.05 (Supplementary Table 5).

In the reverse 2SMR, we identified a strong causal association (*P* < 0.002 or α=0.05/30 CpGs) between cg25536676 (*DHCR24*) and T2D using the Wald ratio (Table 3). For this CpG, we identified opposite direction of association between the causal and the observational analysis. Similar causal effects with T2D were seen at cg25536676 using the IVW regression (Supplementary Table 6), with no evidence of heterogeneity (Cochran’s Q=2.4, *P*=0.30). The Steiger test supported the knowledge that true direction of association at cg25536676 was from DNAm to T2D (Steiger *P*=3.8×10^−147^, R^2^ CpG=0.04, R^2^ T2D=5.3×10^−5^), and not the opposite. Even though we conducted sensitivity MR methods for the association at cg25536676 (Supplementary Table 6), these results may be unreliable due to the small number of mQTL used as instruments (n=3 mQTL).

**Table 3.** Observational and causal estimates of the association between difference in DNAm at five CpGs and prevalent T2D. CpGs shown were observed with the strongest evidence of causality in the reverse 2SMR. Observational estimates were previously obtained from a meta-analysis of EWAS of T2D. Highlighted in bold, association identified with Bonferroni significance at P < 0.002.

| CpG | Chr | Nearest Gene | GWA study | Meta-EWAS of T2D |  | MR Wald ratio |  |  |
| --- | --- | --- | --- | --- | --- | --- | --- | --- |
|  |  |  |  | Effect (95%CI) | P | OR (95%CI) | Effect (95%CI) | P |
| cg25536676 | 1 | *DHCR24* | Mahajan et al. 2014 | -0.008(-0.011,-0.004) | 5.39E-06 | 1.43 (1.15,1.78) | 0.36 (0.14,0.58) | 0.001 |
| cg11851382 | 1 | *PPAP2B* | Wood et al. 2016 | -0.008 (-0.011, -0.005) | 6.42E-06 | 0.29 (0.12,0.67) | -1.24 (-2.09,-0.4) | 0.004 |
| cg10082515 | 7 | *Open sea* | Wood et al. 2016 | -0.013(-0.019,-0.008) | 7.46E-06 | 0.68 (0.52,0.89) | -0.38 (-0.65,-0.11) | 0.005 |
| cg07212837 | 8 | *Open sea* | Mahajan et al. 2014 | 0.006(0.004,0.009) | 3.28E-06 | 1.09 (1.02,1.17) | 0.09 (0.02,0.16) | 0.018 |
| cg16765088 | 15 | *SYNM* | Wood et al. 2016 | -0.011(-0.014,-0.007) | 5.50E-10 | 1.72 (1.02,2.9) | 0.54 (0.02,1.07) | 0.040 |

GWA study: genome-wide association study used to extract information of the genotype-outcome associations. Meta-EWAS of T2D: meta-analysis of epigenome-wide association studies of prevalent T2D (N=3,428) [6]. MR: Mendelian randomization. Associations were considered significant at $P < 0.002$ or $\alpha = 0.05/30$ CpGs analyzed in the reverse 2SMR.

We observed suggestive evidence (unadjusted *P* < 0.05) of a causal effect of DNAm on T2D at other four CpG sites (Table 3). Estimates of the causal and the observational analysis were consistent for most of these associations, except for that at the CpG in *SYNM*. No other MR methods were applied at these four CpGs as they were proxied by a single mQTL.

Overall, results of the Steiger test suggested that true direction of association was the one we were interrogating in the reverse 2SMR for the top CpGs identified in this analysis (Steiger P range 1.2×10^−164^ to 1.5×10^−94^, mean R^2^ for CpGs= 0.03 vs mean R^2^ for T2D= 2.3×10^−5^). An average F-statistic of 20.3 suggested less probability of obtaining biased results due to weak instruments. In Figure 4, we show volcano and forest plots summarizing results of the reverse 2SMR.

**Figure 4.**
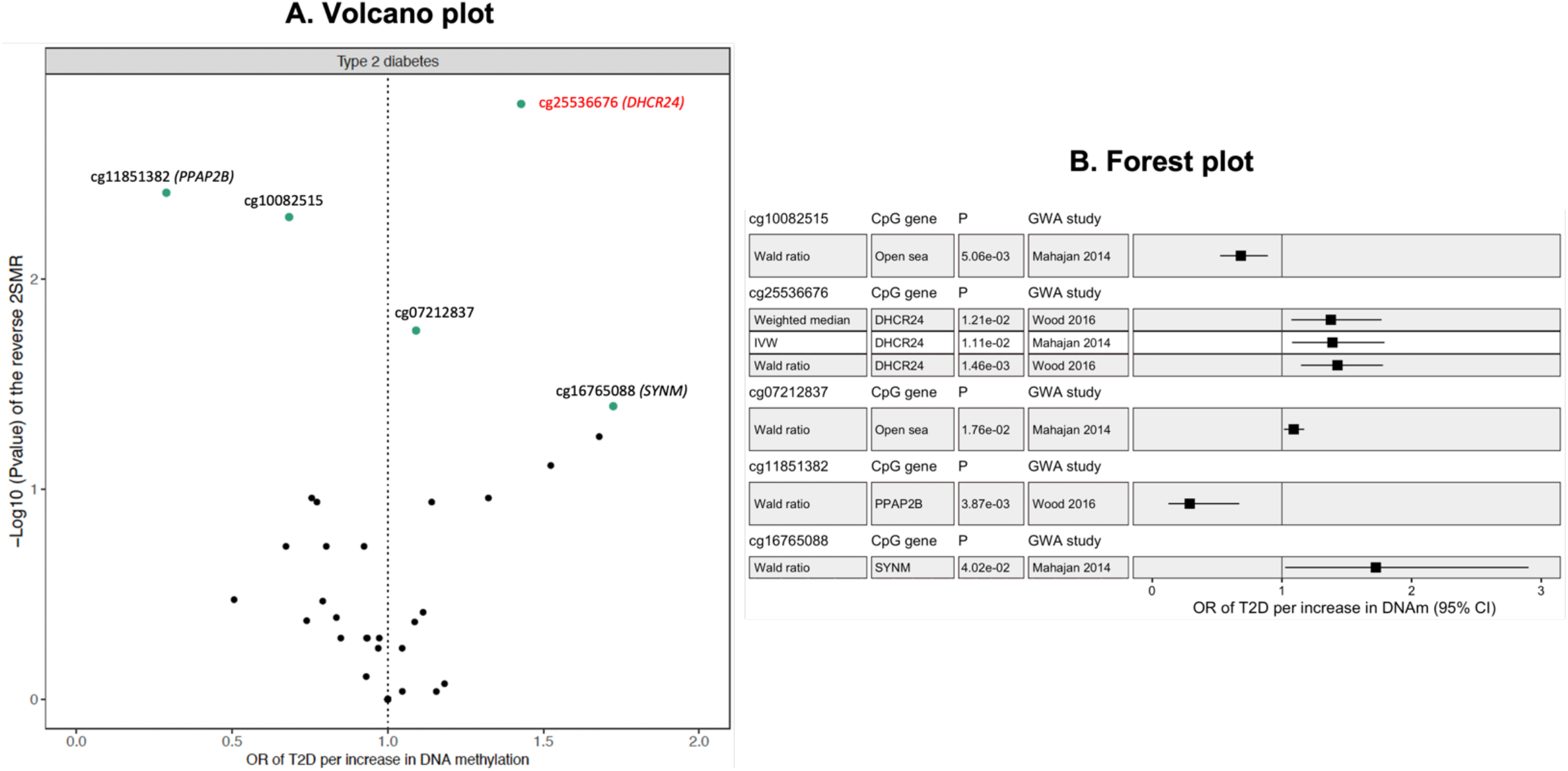
Summarizing evidence of the reverse 2SMR for the association between inverse-normal transformed residuals of DNAm at five meta-EWAS CpGs, and risk of prevalent T2D. CpGs illustrated showed the smallest P-value in the reverse 2SMR. A) Volcano plot showing MR estimates using the Wald ratio and a single SNP as proxy for each CpG analyzed. Highlighted in red is the CpG cg25536676 (DHCR24) identified as causally associated with T2D (P= 0.001). Green dots are CpGs identified with unadjusted P < 0.05 or -Log10(P-value) > 1.3 in the 2SMR. Vertical dashed line represents the line of null associations at OR= 1.0. B) Forest plot showing mean and standard error of the causal estimates at top five CpGs identified in the reverse 2SMR. Results for each CpG are grouped by the MR method applied, which differed only for the CpG cg25536676 (DHCR24) proxied by 3 mQTL. 2SMR: Two-sample Mendelian randomization.

### Bidirectional interrogation of associations identified in the 2SMR

To determine true direction of association with T2D, we summarized bidirectionally causal effects derived from the 2SMR. For the seven top CpGs with bidirectional 2SMR data (2 detected in forward 2SMR and 5 in the reverse 2SMR) (Table 4), we confirmed that true direction of association was the one we originally reported in the single-direction analysis. At cg10082515, we observed the same direction of association between the observational and the bidirectional causal analysis (Table 4). For four of the top six CpGs previously identified in a meta-EWAS of T2D that had bidirectional 2SMR data (CpGs in *TXNIP, HDAC4, SYNM* and *ABCG1*), we saw consistency between the observational and causal effects only at cg00144180 (*HDAC4*). At this CpG, MR results suggested that true direction of association was from DNAm to T2D, similar to what we observed at the CpG cg16765088 in *SYNM* (Supplementary Table 7). For the CpGs cg19693031 (*TXNIP*) and cg06500161 (*ABCG1*), results of the 2SMR were inconclusive (i.e., very large P-values in both directions of the 2SMR), suggesting no evidence of a causal effect of DNAm on T2D. The CpG cg00574958 in *CPT1A*, previously shown as causally associated with fasting glucose [7], was not associated with T2D in the single direction where it was analyzed (forward 2SMR: T2D ⟶ DNAm).

**Table 4.** Bidirectional comparison of MR estimates obtained for CpGs associated with T2D with significance or borderline significance in either direction of the two-sample MR analysis. In addition, showing estimates of the observational analysis (meta-EWAS of T2D).

| CpG | Nearest gene | Meta-EWAS of T2D |  | Forward 2SMR<br>(T2D → DNAm) |  | Reverse 2SMR<br>(DNAm → T2D) |  | Likely causal direction |
| --- | --- | --- | --- | --- | --- | --- | --- | --- |
|  |  | Estimate (95%CI) | P | Estimate (95%CI) | P | Estimate (95%CI) | P |  |
| cg11851382 | *PPAP2B* ** | -0.008(-0.011,-0.004) | 6.42E-06 | 0.03(-0.09,0.15) | 0.603 | -1.24(-2.09,-0.40) | 0.004 | DNAm → T2D |
| cg20812370 | *PBX1* * | -0.007(-0.009,-0.004) | 7.40E-07 | -0.18(-0.30,-0.07) | 0.002 | --- | --- | T2D → DNAm |
| cg25536676 | *DHCR24* ** | -0.008(-0.011,-0.004) | 5.39E-06 | 0.04(-0.08,0.16) | 0.490 | 0.36(0.14,0.58) | 0.001 | DNAm → T2D |
| cg20456243 | *SPEG* * | -0.007(-0.011,-0.004) | 9.99E-06 | -0.07(-0.19,0.06) | 0.304 | 0.08(-0.12,0.29) | 0.429 | T2D → DNAm |
| cg10082515 | *Open sea* ** | -0.013(-0.019,-0.008) | 7.46E-06 | -0.05(-0.16,0.07) | 0.443 | -0.38(-0.65,-0.11) | 0.005 | DNAm → T2D |
| cg07212837 | *Open sea* ** | 0.006(0.004,0.009) | 3.28E-06 | -0.07(-0.2,0.06) | 0.304 | 0.09(0.02,0.16) | 0.018 | DNAm → T2D |
| cg24686009 | *RAP1B* * | -0.002(-0.003,-0.001) | 1.19E-06 | 0.004(-0.11,0.12) | 0.942 | --- | --- | Inconclusive† |
| cg16765088 | *SYNM* ** | -0.011(-0.014,-0.007) | 5.50E-10 | -0.08(-0.20,0.04) | 0.188 | 0.54(0.02,1.07) | 0.04 | DNAm → T2D |
| cg01577083 | *Open sea* * | -0.011(-0.016,-0.006) | 7.93E-06 | -0.15(-0.28,-0.02) | 0.023 | 0.00(-0.32,0.32) | 1.00 | T2D → DNAm |
| cg26766064 | *MIR657* * | -0.007(-0.009,-0.004) | 5.17E-06 | -0.10(-0.22,0.02) | 0.096 | --- | --- | T2D → DNAm |

Meta-EWAS of T2D: meta-analysis of epigenome-wide association studies of prevalent T2D (N=3,428). 2SMR: Two-sample Mendelian randomization. DNAm: DNA methylation. Associations were considered significant at $P < 1.33 \times 10^{-7}$ in the meta-EWAS of T2D, $P < 0.001$ in the forward 2SMR, and $P < 0.002$ in the reverse 2SMR analysis. \* CpGs identified with borderline significance in the forward 2SMR. \*\* CpGs identified with significance or borderline significance in the reverse 2SMR. †Inconclusive causal direction for CpGs were 2SMR data was available only in one direction and $P > 0.1$ , or if 2SMR data was present bidirectionally but in both cases $P$ was $> 0.1$ .

### *In silico* interrogation of the functional role of meta-EWAS CpGs analyzed in a bidirectional 2SMR

We identified enrichment for eQTMs among CpGs analyzed in the 2SMR (OR= 5.4, 95% CI= 2.7-11.1, *P*=1.5×10^−7^). Nine eQTMs were found among CpGs with evidence of association with T2D in the forward (T2D⟶ DNAm; cg12593793, cg20456243 (*SPEG*), cg11024682 (*SREBF1*) and cg27037013) or the reverse 2SMR (DNAm ⟶T2D; cg18181703 (*SOCS3*)), also among those with inconclusive direction of association (cg14476101 (*PHGDH*), cg19693031 (*TXNIP*), cg00574958 (*CPT1A*), cg06500161 (*ABCG1*)) (Table 5). We also interrogated enrichment of meta-EWAS CpGs for traits included in the EWAS catalog, categorizing CpGs into three groups following evidence from the bidirectional 2SMR (see Methods). Common to all groups was the higher enrichment for ancestry/ethnicity, anthropometric and cardiometabolic traits. Unique to the first group (T2D causal of DNAm), was the enrichment for neurological and perinatal traits, while unique to the second group (DNAm causal of T2D), was the enrichment for lipid lipoproteins, alcohol, and metabolites. In the third group of CpGs with “inconclusive direction of association”, we found enrichment for all the above traits in addition to age and tissue. Forest plots summarizing evidence of the enrichment analysis are presented in the Supplementary Figure 3.

**Table 5.** Look up of expression quantitative trait methylation sites (eQTMs) among meta-EWAS CpGs included in a bidirectional two-sample MR analysis. Association estimates as retrieved from the BIOS QTL browser[37].

| CpG | Transcript | Chr | CpG position | Transcript position | Gene | Beta (SE) | P | FDR |
| --- | --- | --- | --- | --- | --- | --- | --- | --- |
| cg14476101 | ENSG00000092621 | 1 | 120255944 | 1.2E+08 | *PHGDH* | 0.34 (0.04) | 2.1E-55 | 0.0E+00 |
| cg19693031 | ENSG00000117289 | 1 | 145441552 | 1.45E+08 | *TXNIP* | -0.12 (0.04) | 7.1E-08 | 9.3E-06 |
| cg12593793 | ENSG00000160789 | 1 | 156074135 | 1.56E+08 | *LMNA* | -0.21 (0.04) | 5.1E-18 | 0.0E+00 |
| cg20456243 | ENSG00000072195 | 2 | 220352428 | 2.2E+08 | *SPEG* | 0.08 (0.04) | 2.4E-05 | 6.0E-03 |
| cg00574958 | ENSG00000110090 | 11 | 68607622 | 68611878 | *CPT1A* | -0.17 (0.04) | 3.1E-20 | 0.0E+00 |
| cg11024682 | ENSG00000072310 | 17 | 17730046 | 17740325 | *SREBF1* | -0.19 (0.04) | 4.5E-15 | 0.0E+00 |
| cg18181703 | ENSG00000184557 | 17 | 76354573 | 76356158 | *SOCS3* | 0.13 (0.04) | 1.1E-06 | 3.3E-04 |
| cg27037013 | ENSG00000237945 | 21 | 35320619 | 35287838 | *LINC00649* | 0.14 (0.04) | 4.8E-07 | 1.3E-04 |
| cg06500161 | ENSG00000160179 | 21 | 43656587 | 43619799 | *ABCG1* | -0.32 (0.04) | 2.2E-37 | 0.0E+00 |

## DISCUSSION

This study conducted a bidirectional two-sample MR to investigate causality in the association between prevalent T2D and DNAm at 58 CpGs previously identified in a meta-EWAS of T2D among Europeans. In the forward 2SMR, where we examined if T2D was causal of differences in DNAm, we interrogated causality at all the 58 meta-EWAS CpGs. No tests passed our p-value threshold. CpG cg20812370 (*PBX1*) had the smallest p-value (p= 0.002) with similar results after implementing sensitivity analyses. For the reverse 2SMR, where DNAm was the exposure, we tested causality at 30/58 meta-EWAS CpGs. We demonstrated that elevated DNAm at cg25536676 (*DHCR24*) was causally associated with increased risk of T2D, which is inconsistent with previous observational evidence. By assessing MR and observational estimates, we attempted to infer the likely causal direction for the 30 CpGs analyzed bidirectionally. Considering directionality consistency with estimates of the observational analysis and smallest P-value in the 2SMR (*P* <0.1), we concluded that for 15 (50%) of the 30 CpGs DNAm was likely causal of T2D, for 10 (33%) of them T2D was causal of DNAm, and for the remaining 5 (17%) CpGs results were inconclusive. *In silico* analyses showed that some of the CpGs analyzed correspond to eQTMs, and that CpGs may have enrichment for specific traits dependent on the direction of causality predicted by the 2SMR.

### Forward 2SMR findings

We did not identify any associations in the forward 2SMR. This may have occurred because there were power limitations with the data available for the analysis of the genotype-outcome association (i.e., ALSPAC-ARIES vs GoDMC). Despite previous nominal and robust associations of *CPT1A* methylation with T2D [11] and fasting glucose [7], we could not confirm this association in our 2SMR analysis.

### Reverse 2SMR findings

Our results of the bidirectional 2SMR suggested that most of the associations investigated may occur due to differences in DNAm influencing the risk of T2D, rather than the opposite, with the most robust evidence obtained at cg25536676 (*DHCR24*). We found that increased levels of inverse normal-transformed residuals of DNAm at this CpG were associated with 43% higher risk of T2D, and this result was consistent among the sensitivity methods applied, but it was opposite to results of the observational analysis [6]. No evidence of heterogeneity or pleiotropic effects were seen for the association at cg25536676 (*DHCR24*). DNAm at cg25536676 has been previously found in association with total cholesterol [40] in whole blood, and with perinatal traits like gestational age [41] and birthweight [42] in cord blood.

Other CpGs in the region of *DHCR24* (cg17901584 and cg27168858) have been identified in association with HDL-cholesterol [40], triglycerides [43], fasting insulin [44, 45], BMI, glycated hemoglobin A1c or HbA1c, incident T2D [45], statin use [46], waist circumference (WC) [47], and LDL-cholesterol [48]. Both CpGs in *DHCR24* were positively associated with HDL-cholesterol and LDL-cholesterol, and cg17901584 was negatively associated with the remaining traits. Our results of the MR at cg25536676 in relation to prevalent T2D, are directionally consistent with the associations identified with the lipid traits at nearby CpGs in *DHCR24*. Considering DNAm at *DHCR24* as the outcome, one study showed evidence of causality in the association with LDL-cholesterol using a stepwise MR approach [48], while only some evidence of causality was identified with BMI [45]. Because obesity and dysregulation in the metabolism of lipids are well-known hallmarks of T2D [49], and because *DHCR24* encodes for an enzyme related with the metabolism of cholesterol (3-hydroxysterol-24 reductase) [40], an association between methylation at *DHCR24* and T2D is very plausible. Thus, DNAm at our CpG cg25536676 in *DHCR24* may act as a causal mediator in the association between known risk factors and T2D. However, a formal two-step two-sample MR analysis is required to prove this hypothesis. Taken together, our results suggest that cg25536676 may be a new DNAm target for the early detection and treatment of T2D.

### Functional analysis

The *in-silico* analysis using data from previous EWAS showed that CpGs detected as potentially causal of T2D may be associated with lipid lipoproteins, metabolites, anthropometric and cardiometabolic traits, all of them involved in the onset of T2D. For comparison, CpGs that were likely to be secondary to the effects of T2D (in *PBX1, SPEG, MIR657* and cg01577083) based on results of the 2SMR, were enriched for perinatal and neurological traits, in addition to cardiometabolic and anthropometric traits. Although not confirmatory, these results indicate that DNAm may act in the development and progression of T2D through different mechanisms related with specific stages in the pathophysiology of the disease. We hypothesize that the observed enrichment of ancestry/ethnicity among T2D CpG sites identified in Europeans, may highlight common confounders or exposures (e.g., differences in cell composition or T2D risk factors) between populations [50].

### Strengths and limitations

Our study has several strengths. First, we were able to leverage large datasets to extract genetic associations with T2D and DNAm. Second, we analyzed causality bidirectionally to disentangle true direction of association and rule out reverse causation at CpGs previously detected in the context of prevalent T2D. We also conducted sensitivity analyses to validate assumptions of the MR analysis, and we demonstrated that in each direction of the analysis we were unlikely to be affected by weak instrument bias. In addition, we run *in-silico* functional analyses to facilitate biological interpretation of MR findings. Study limitations included the use of a smaller outcome sample in the forward 2SMR. Furthermore, in the reverse MR analysis (DNAm ⟶ T2D), we had few proxies to predict DNAm at the CpGs of interest (∼ 1 mQTL/CpG). This prevented us from obtaining stronger evidence of causality and conducting sensitivity MR analyses. We also did not account for potential overlap between the exposure and outcome samples in the reverse 2SMR (up to 9.2% overlap), which could have biased our results. Lastly, observational evidence used to conduct the 2SMR was focused on Europeans, which limits the generalizability of our MR findings to other populations. Thus, future studies will benefit from the use of multi-ethnic cohorts throughout the observational and causal inference stages of the analysis. Likewise, a two-step 2SMR may be required to investigate the role of DNAm as a potentially causal mediator between known risk factors and T2D.

## CONCLUSIONS

In this study we assessed causality between DNA methylation and T2D in a bidirectional 2SMR framework at CpG sites identified from a meta-EWAS of prevalent disease. Due to the strength of the genetic associations retrieved, we had more power to identify causality from DNAm to T2D. We demonstrated that a CpG in *DHCR24*, a gene related with the metabolism of lipids, was causally associated with T2D. Validation of associations identified with causality or suggestive evidence of causality in this study may benefit from the use of larger and better powered samples to extract genetic associations, especially when DNAm is the exposure.

## Data Availability

ALSPAC data used for this submission will be made available on request to the ALSPAC
executive committee. The ALSPAC data management plan
(available here: www.bristol.ac.uk/alspac/researchers/access/) describes in detail the policy
regarding data sharing, which is through a system of managed open access.

https://diagram-consortium.org

https://github.com/hannahe/collapse_EWAS_catalog_phenotypes/blob/9b65be66399d0c1d2fd71c2003dbf58e4e5b62ff/functional_analysis_regroup_EWAS_catalogue_phenotypes.R

https://genenetwork.nl/biosqtlbrowser/

http://www.ariesepigenomics.org.uk/

http://www.godmc.org.uk/

## AUTHORS CONTRIBUTIONS

DJQ, HRE, GS and CLR contributed to the conceptualization of the study. DJQ led the data analysis and wrote the manuscript with HRE. ES provided intellectual input on the 2SMR methods utilized. All authors contributed to discussion, provided critical input to the project and drafting of the manuscript.

## ACKNOWLEDGMENTS

All authors work in the Medical Research Council Integrative Epidemiology Unit at the University of Bristol, which is supported by the Medical Research Council and the University of Bristol (MC_UU_00011/5 and MC_UU_00011/1).

ALSPAC: We are extremely grateful to all the families who took part in this study, the midwives for their help in recruiting them, and the whole ALSPAC team, which includes interviewers, computer and laboratory technicians, clerical workers, research scientists, volunteers, managers, receptionists and nurses.

The UK Medical Research Council and Wellcome (Grant ref: 217065/Z/19/Z) and the University of Bristol provide core support for ALSPAC. A comprehensive list of grants funding is available on the ALSPAC website (http://www.bristol.ac.uk/alspac/external/documents/grant-acknowledgements.pdf). This publication is the work of the authors and HRE will serve as guarantor for the contents of this paper

## ETHICAL DISCLOSURE

Ethical approval for ALSPAC and its sub study ARIES was obtained from the ALSPAC Ethics and Law Committee and the Local Research Ethics Committees. Consent for biological samples has been collected in accordance with the Human Tissue Act (2004).

## DATA ACCESS STATEMENT

ALSPAC data used for this submission will be made available on request to the ALSPAC executive committee. The ALSPAC data management plan (available here: www.bristol.ac.uk/alspac/researchers/access/) describes in detail the policy regarding data sharing, which is through a system of managed open access.

