## Supplementary material for "Investigating causality in the association between DNA methylation and prevalent T2D using a bidirectional two-sample Mendelian Randomization"

### S1. ALSPAC ARIES

Pregnant women resident in Avon, UK with expected dates of delivery 1st April 1991 to 31st December 1992 were invited to take part in the study. The initial number of pregnancies enrolled is 14,541 (for these at least one questionnaire has been returned or a "Children in Focus" clinic had been attended by 19/07/99). Of these initial pregnancies, there was a total of 14,676 foetuses, resulting in 14,062 live births and 13,988 children who were alive at 1 year of age [1, 2]. When the oldest children were approximately 7 years of age, an attempt was made to bolster the initial sample with eligible cases who had failed to join the study originally. As a result, when considering variables collected from the age of seven onwards (and potentially abstracted from obstetric notes) there are data available for more than the 14,541 pregnancies mentioned above. The number of new pregnancies not in the initial sample (known as Phase I enrolment) that are currently represented on the built files and reflecting enrolment status at the age of 24 is 913 (456, 262 and 195 recruited during Phases II, III and IV respectively), resulting in an additional 913 children being enrolled. The phases of enrolment are described in more detail in the cohort profile paper and its update [1, 2]. The total sample size for analyses using any data collected after the age of seven is therefore 15,454 pregnancies, resulting in 15,589 foetuses. Of these 14,901 were alive at 1 year of age. A 10% sample of the ALSPAC cohort, known as the Children in Focus (CiF) group, attended clinics at the University of Bristol at various time intervals between 4 to 61 months of age. The CiF group were chosen at random from the last 6 months of ALSPAC births (1432 families attended at least one clinic) [1, 2]. Excluded were those mothers who had moved out of the area or were lost to follow-up, and those partaking in another study of infant development in Avon. In our MR study of T2D, we considered a subsample of 867 mothers and 385 fathers from the main ALSPAC cohort with genetic and epigenetic (DNA methylation) data to conduct analyses.

Please note that the study website contains details of all the data that is available through a fully searchable data dictionary and variable search tool (http://www.bristol.ac.uk/alspac/researchers/our-data/)

Ethical approval for the study was obtained from the ALSPAC Ethics and Law Committee and the Local Research Ethics Committees. Consent for biological samples has been collected in accordance with the Human Tissue Act (2004).

**S2. Extraction of genotype data from ALSPAC-ARIES samples**

Genotyping was completed prior to this project. Thus, for middle-aged adults in ALSPAC, genetic data for 148 T2D SNPs was extracted from the ALSPAC GWAS database [1] using the latest genetic imputation datasets available (mothers: release 2015-10-30, fathers: release 2016-11-22). Because imputation was done independently for mothers and fathers in ALSPAC, both datasets were merged retaining the first ten genetic principal components (PCs) to adjust for genetic structure in downstream analyses. Detail of the method applied for genotyping and imputation of genetic data in mothers in ALSPAC has been described elsewhere [1, 3]. Briefly, genetic data was generated using the Illumina Infinium Human660W-Quad BeadChip array v1.0, and the Illumina GenomeStudio software for genotyping calling (genome build 37). Quality control (QC) measures applied included removal of SNPs with minor allele frequency (MAF) < 0.01, Hardy-Weinberg equilibrium P-value < $10^{-6}$, and missing genotyping rate > 0.05. In addition, samples with indeterminate X chromosome heterozygosity, genotyping missingness higher than 5%, and evidence of population stratification, were excluded. Data was imputed to the 1,000 Genomes (phase 1, version 3, http://www.internationalgenome.org/about) using Impute2 version 2.2.2, retaining samples with MAF > 0.01 and calling rate > 80%.

Genetic data for the fathers was genotyped using the Illumina HumanCoreExome BeadChip array, and the Illumina GenomeStudio software for variant calling. QC measures included removing SNPs with Hardy-Weinberg equilibrium P-value < $10^{-7}$, SNPs failing GenomeStudio QC, and SNPs that were duplicated. Samples with gender mismatch, high or low heterozygosity, genotyping missingness > 5%, contamination, and of non-European origin, were excluded. Before imputation, genotype data for 3,074 fathers (some of them related) was furtherly controlled for variants not included in the 1,000 Genomes, monomorphic SNPs and duplicated sites. Imputation was performed as in the mother's dataset. Data from the imputation was retained if MAF > 0.01 and calling rate > 80%.

### S3. Quality control of genetic data in ALSPAC-ARIES

Imputed genetic data extracted for middle-aged adults in ALSPAC-ARIES was QC before the analysis. Imputed genotype data was available for 6,102,837 SNPs after merging the genetic datasets of mothers and fathers in ALSPAC using a consensus method in Plink. Genotyping rate for these variants was 0.98, and further inspection of the data included plotting imputation quality scores against the MAF to verify that variants remaining in the dataset had MAF > 0.01, and high imputation rate (info > 0.86). Additional QC for the SNPs included removing those with MAF < 0.01, calling rate < 0.8, missing genotyping rate > 0.05, and Hardy-Weinberg equilibrium p-value < $10^{-6}$. Samples were excluded if cryptic relatedness between pairwise comparisons was higher than 0.125, and if missing genotyping rate was > 0.05. Twenty PCs were generated for the genotype data of unrelated samples using a list of 26,873 independent SNPs identified in the HapMap3 project with Linkage Disequilibrium or LD < 0.1 and MAF > 0.2. None of the samples were identified as an outlier based on genetic variation measured by the PCs. Twenty genetic PCs were used as covariates in processing of the DNA methylation data. In total, 1,243 middle-aged adults and approximately 5.3 million SNPs remained in the dataset after QC. Total genotyping rate was 0.99, average missing genotyping rate was 0.01, and correlation between observed and expected allele frequencies in the 1,000 Genomes was 0.99.

### S4. Quality control of DNA methylation data in ALSPAC-ARIES

Genome-wide DNA methylation was available for 482,015 probes, including probes in sex chromosomes and probes reported in the Naeem list [4]. Methylation data was initially QC for outliers, which were identified as samples with methylation levels ten standard deviations higher than the mean of the probe. Outliers were detected after three iterations and replaced by the mean of the probe.

Normalization of the methylation data was applied to avoid false positives in the SNP-CpG analysis, and to minimize the amount of non-genetic residual variation remaining in the data. To achieve this, methylation was first inverse-normal transformed, and then regressed against covariates and twenty genetic PCs previously generated (see section S3). We obtained residuals of DNAm using a regression model adjusted for age, sex, batch effects (i.e., bisulphite conversion plate), predicted counts for seven white-blood cells (Houseman method), and predicted levels of smoking. Predicted levels of smoking were estimated using data from two DNAm scores developed by Zeilinger *et al.* [5] and by Elliott *et al.* [6]. Because only complete data was required in the dataset of covariates, missing data for age in a subset of males was replaced using predictive mean matching imputation method in the R package MICE [7]. Covariates and genetic PCs were regarded as fixed effects in the regression model. Residuals of this analysis were then regressed against methylation PCs estimated to account for residual confounding. Methylation PCs were generated using the 20,000 most variable autosomal probes, and they were tested for association with the genotype (i.e., 62 T2D SNPs) using a linear regression model in the MatrixeQTL R package. Methylation PCs associated with the genotype at P-value $< 10^{-7}$ were excluded. In total, the first thirteen non-genetic methylation PCs were retained for further analyses. Residuals of the regression between adjusted methylation values at 482,015 probes and methylation PCs, were used in the SNP-CpG analysis. The linear regression model used for analysis in MatrixeQTL was the following:

$$\text{CpGn} = \alpha + \Sigma_k \beta_k \cdot \text{covariate}_k + \gamma \cdot \text{genotype\_additive}$$

Where $\beta_k$ and $\gamma$ are the effect of covariates and the genotype (i.e., per effect allele) on variation in DNA methylation, respectively.

**S5. Selecting genetic proxies for meta-EWAS CpGs in GoDMC**

To assess if increased DNAm at selected meta-EWAS CpGs was causal of T2D risk, we searched for genetic proxies for 58 meta-EWAS of T2D CpGs in the GoDMC consortium. GoDMC is currently the biggest consortium for the study of the genetics of DNAm variation, comprising 32,851 samples across 36 contributing cohorts [8]. Samples included in GoDMC were mostly of European ancestry, with mean age of 55.6 years. In the discovery stage, around 248,607 independent *cis*-mQTL (SNP < 1 Mb from CpG site) and 23,117 *trans*-mQTL (SNP > 1 Mb from CpG site) were reported using a P-threshold $< 10^{-8}$ and $P < 10^{-14}$, respectively [8]. Additional information about GoDMC and their study design, can be found elsewhere [8].

Of the 58 T2D CpGs of interest, 31 had at least one mQTL reported in GoDMC. Of these 31 CpGs, 22 had one mQTL, 8 CpGs (in *HDAC4*, *ITIH1*, *PHGDH*, *ROBO1*, *SMYD5*, cg00896068, cg27037013 and cg27115863) had two mQTL, and the CpG in *DHCR24* had three mQTL reported in GoDMC. Association estimates between the mQTL SNPs and T2D were retrieved from two previously published GWAS in T2D [9, 10] included in the MR-Base database. From these two studies, we were able to successfully extract genetic association data with T2D for 39 mQTL SNPs typing 30 meta-EWAS CpGs after data harmonization in MR-Base. We excluded the mQTL SNP rs74623153 (typing cg11983038) without GWAS information, and rs9976794 (typing cg27037013) that was a palindromic SNP with intermediate allele frequencies.

**S6. Conducting Mendelian Randomization analysis using MR-Base**

We implemented tools available in the TwoSampleMR (version 0.5.6), MRInstruments (version 0.3.2) and MendelianRandomization (version 0.5.1) R packages to conduct causal analyses [11, 12]. We applied *clumping* to select only independent instruments (LD $r^2 < 0.01$) for T2D and DNAm. For the MR analysis, we used default parameters in MR-Base and selected as our main findings results of the inverse-variance weighted (IVW) regression using a random-effect model, or those from a Wald ratio when the number of instruments available was < 2 SNPs. When there were > 2 instruments, we included three sensitivity analyses for comparison with results of the IVW: the MR-Egger regression to account for horizontal pleiotropy in the effect of proxy SNPs on the outcome [12], and the weighted median and weighted mode regressions to account for potential invalid instruments [11, 12]. Evidence of directional pleiotropy was considered if the intercept of the MR-Egger regression had a $P < 0.001$ ($\alpha = 0.05/62$ T2D SNPs). We applied correction for multiple testing in results of the MR analysis using an $\alpha = 0.05/58$ CpGs or $P < 1.0\times10^{-3}$ for the forward MR (T2D → 58 meta-EWAS CpGs), or an $\alpha = 0.05/30$ CpGs or $P < 2.0\times10^{-3}$ for the reverse MR (30 successfully proxied meta-EWAS CpGs → T2D). We deemed as suggestive of causality associations with an uncorrected $P < 0.05$.

We measured SNP heterogeneity using the Cochran's Q estimate derived from the IVW, and the Rucker's Q estimate obtained from the MR-Egger regression. Heterogeneity was significant if Q > 70 and $P < 0.001$. We used the Steiger test to verify that true direction of association was the one specified in the analysis. This analysis may perform better when using continuous exposures (i.e., DNAm as opposed to T2D). Lastly, we implemented a leave-one-out analysis using the IVW method to identify potential outlier SNPs [11]. Strength of the instruments was estimated in each direction of the two-sample MR using the F-statistic reported by the IVW, the $I^2$ statistic ($I_{GX}^2$) reported by the MR-Egger regression, or it was manually calculated as the beta-coefficient/standard error for the SNP-exposure association. We disregarded presence of weak instrument bias in MR findings if the F-statistic > 10 or if $I_{GX}^2 \sim 1.0$. When possible, different graphical methods, like scatter plots, funnel plots, forest plots and volcano plots, were used to represent results of the MR analysis and validate MR assumptions.

**S7. Results of sensitivity analyses implemented in the forward 2SMR (T2D → DNAm)**
When we applied additional MR methods with less stringent assumptions about the validity of the instruments like the weighted median and weighted mode analyses, they showed same direction of association and similar P-value as results of the IVW at top two CpGs identified in the forward 2SMR, cg20812370 (*PBX1*) and cg01577083. However, absolute effect estimates were slightly larger when using these other methods. Even though we found no evidence of pleiotropy in the association at cg20812370 (*PBX1*) [Cochran's Q estimate 52.9, *P*= 0.76], we observed some asymmetry in the funnel plot for this site attributed to two outlier SNPs, rs319598 and rs1359790, which in the forest plot were also seen with a more negative effect in cg20812370 (*PBX1*) compared to the rest of the SNPs (Supplementary Figure 1). We re-ran the analysis for this CpG excluding the two outlier SNPs, and we observed a less negative effect estimate but a larger P-value this time around (*P* = 0.002 with and *P* = 0.01 without outlier SNPs). Applying a leave-one-out analysis to investigate the effect of influential SNPs on the outcome (DNAm), we did not find evidence that any single T2D SNP was independently driving the total causal estimate at cg20812370 (*PBX1*) or at cg01577083. The Steiger test suggested that true direction of association at cg01577083 was from DNAm to T2D, and not the direction here analyzed (Steiger $P$=5.5x10$^{-5}$, $R^2$ for T2D = 0.03 < $R^2$ for cg01577083 = 0.07). Summary plots of the forward 2SMR for cg20812370 (*PBX1*) and cg01577083 can be found in the Supplementary Figures 1-2.

**Supplementary Table 1. Four GWAS of T2D reported in the DIAGRAM consortium and used to extract genetic proxies for T2D in the forward two-sample MR analysis (T2D → meta-EWAS CpGs).**

| SNPs | Data source | Population | Discovery sample | Replication sample | Notes |
|---|---|---|---|---|---|
| 60 | Morris *et al.* 2012<br><br>PMID: 22885922 | Europeans | 121,171 cases and 56,862 controls | 22,669 cases and 58,119 controls | Genotyping method was the Metabochip array. Meta-analysis of GWAS adjusted for genomic inflation |
| | | Pakistani (PROMIS) | | 1,178 cases and 2,472 controls | |
| 34 | Mahajan *et al.* 2014<br><br>PMID: 24509480 | Europeans | 12,171 cases and 56,862 controls | 21,491 cases and 55,647 controls | Ancestry-specific GWAS corrected for study-specific covariates and genomic inflation. Trans-ethnic meta-analysis corrected for genomic inflation. |
| | | South Asians | 6,952 cases and 11,865 controls | | |
| | | East Asians | 5,561 cases and 14,458 controls | | |
| | | Mexicans and Mexican Americans | 1,804 cases and 779 controls | | |
| 40 | Gaulton *et al.* 2015 PMID: 26551672 | Europeans | 27,206 cases and 57,574 controls | Not reported | Fine mapping of 39 established genetic loci in T2D |
| 14 | Fuchsberger *et al.* 2016<br><br>PMID: 27398621 | Europeans<br>East Asians<br>South Asians<br>African American Hispanics | 11,645 cases and 32,769 controls | Not reported | |

**Supplementary Table 2. Association estimates of 62 SNPs extracted from the DIAGRAM consortium and used as genetic proxies for T2D in the forward two-sample MR analysis (T2D → meta-EWAS CpGs). Associations are ordered by P-value (from smallest to largest).**

| SNP | Chr | Mapped gene | Estimate | SE | EA | OA | EAF | P | N |
|---|---|---|---|---|---|---|---|---|---|
| rs7903146 | 10 | *TCF7L2* | 0.33 | 0.02 | T | C | 0.26 | 1.20E-139 | 144,178 |
| rs10811660 | 9 | *Unannotated* | 0.24 | 0.02 | G | A | 0.83 | 1.10E-61 | 219,582 |
| rs35261542 | 6 | *CDKAL1* | 0.16 | 0.01 | A | C | 0.28 | 1.50E-50 | 219,582 |
| rs35510946 | 3 | *IGF2BP2* | 0.13 | 0.01 | A | G | 0.3 | 1.10E-39 | 219,582 |
| rs11187140 | 10 | *Unannotated* | 0.11 | 0.01 | G | A | 0.63 | 1.50E-31 | 219,582 |
| rs13266634 | 8 | *SLC30A8* | 0.11 | 0.01 | C | T | 0.68 | 5.00E-28 | 219,582 |
| rs1513272 | 7 | *JAZF1* | 0.1 | 0.01 | C | T | 0.52 | 7.80E-25 | 219,582 |
| rs9936385 | 16 | *FTO* | 0.12 | 0.02 | C | T | 0.40 | 2.60E-23 | 144,178 |
| rs11712037 | 3 | *PPARG* | 0.13 | 0.02 | C | G | 0.86 | 1.70E-20 | 219,582 |
| rs2972156 | 2 | *Unannotated* | 0.09 | 0.01 | G | C | 0.62 | 4.20E-20 | 219,582 |
| rs11257658 | 10 | *Unannotated* | 0.09 | 0.01 | A | G | 0.22 | 1.20E-15 | 219,582 |
| rs72999033 | 19 | *HAPLN4* | 0.15 | 0.02 | T | C | 0.07 | 1.80E-15 | 219,582 |
| rs7607980 | 2 | *COBLL1* | 0.14 | 0.02 | T | C | 0.88 | 8.30E-15 | 92,794 |
| rs6813195 | 4 | *RPS3AP18; RPS14P6* | 0.08 | 0.01 | C | T | 0.73 | 4.10E-14 | 161,639 |
| rs77981966 | 2 | *THADA* | 0.15 | 0.02 | C | T | 0.93 | 4.10E-14 | 219,582 |
| rs11717195 | 3 | *ADCY5* | 0.1 | 0.02 | T | C | 0.78 | 6.50E-14 | 149,821 |
| rs340874 | 1 | *PROX1* | 0.07 | 0.01 | C | T | 0.52 | 5.10E-13 | 219,582 |
| rs7732130 | 5 | *ZBED3-AS1* | 0.08 | 0.01 | G | A | 0.28 | 2.40E-12 | 219,582 |
| rs17676309 | 3 | *ADAMTS9-AS2; MIR548A2* | 0.07 | 0.01 | C | T | 0.59 | 2.80E-12 | 219,582 |
| rs1387153 | 11 | *MTNR1B* | 0.09 | 0.02 | T | C | 0.29 | 1.60E-11 | 144,178 |
| rs2583941 | 12 | *RPSAP52* | 0.1 | 0.02 | A | G | 0.09 | 1.60E-11 | 219,582 |
| rs10276674 | 7 | *DGKB* | 0.08 | 0.01 | C | T | 0.18 | 2.80E-11 | 219,582 |
| rs3803563 | 15 | *PRC1* | 0.08 | 0.01 | A | C | 0.18 | 5.60E-11 | 219,582 |
| rs12571751 | 10 | *ZMIZ1* | 0.08 | 0.01 | A | G | 0.51 | 1.00E-10 | 149,821 |
| rs878521 | 7 | *YKT6; CAMK2B* | 0.07 | 0.01 | A | G | 0.24 | 1.30E-10 | 219,582 |
| rs1552224 | 11 | *ARAP1* | 0.1 | 0.02 | A | C | 0.83 | 1.80E-10 | 144,178 |
| rs516946 | 8 | *ANK1; MIR486* | 0.09 | 0.02 | C | T | 0.77 | 2.50E-10 | 149,821 |
| rs35720761 | 2 | *THADA* | 0.11 | 0.02 | T | C | 0.89 | 3.30E-10 | 92,794 |
| rs780094 | 2 | *GCKR* | 0.06 | 0.01 | C | T | 0.61 | 3.40E-10 | 219,582 |
| rs243020 | 2 | *Unannotated* | 0.06 | 0.01 | G | A | 0.46 | 5.50E-10 | 219,582 |
| rs35658696 | 5 | *PAM* | 0.16 | 0.03 | A | G | 0.96 | 5.70E-10 | 92,794 |
| rs10842994 | 12 | *KLHL42; PTHLH* | 0.1 | 0.02 | C | T | 0.8 | 6.10E-10 | 149,821 |
| rs5215 | 11 | *KCNJ11* | 0.07 | 0.01 | C | T | 0.39 | 8.50E-10 | 149,821 |
| rs1974620 | 7 | *Unannotated* | 0.06 | 0.01 | T | C | 0.52 | 1.00E-09 | 219,582 |
| rs1496653 | 3 | *UBE2E2; MIR548AC* | 0.09 | 0.02 | A | G | 0.79 | 3.60E-09 | 149,821 |

**Supplementary Table 2. (*Continued*)**

| SNP | Chr | Mapped gene | Estimate | SE | EA | OA | EAF | P | N |
|---|---|---|---|---|---|---|---|---|---|
| rs17106184 | 1 | *FAF1* | 0.1 | 0.02 | G | A | 0.91 | 4.10E-09 | 161,585 |
| rs7177055 | 15 | *Unannotated* | 0.08 | 0.01 | A | G | 0.72 | 4.60E-09 | 149,821 |
| rs2796441 | 9 | *LOC101927502* | 0.07 | 0.01 | G | A | 0.63 | 5.40E-09 | 147,724 |
| rs41278853 | 22 | *MTMR3* | 0.13 | 0.03 | A | G | 0.89 | 5.60E-09 | 92,794 |
| rs6808574 | 3 | *BCL6; LPP-AS2* | 0.07 | 0.01 | C | T | 0.6 | 5.80E-09 | 140,087 |
| rs7955901 | 12 | *Unannotated* | 0.07 | 0.01 | C | T | 0.42 | 6.50E-09 | 144,178 |
| rs702634 | 5 | *ARL15* | 0.06 | 0.01 | A | G | 0.71 | 6.90E-09 | 154,797 |
| rs4275659 | 12 | *ABCB9* | 0.06 | 0.01 | C | T | 0.67 | 9.50E-09 | 161,459 |
| rs12970134 | 18 | *RPS3AP49; MC4R* | 0.08 | 0.02 | A | G | 0.26 | 1.20E-08 | 138,946 |
| rs1359790 | 13 | *LINC01080; SPRY2* | 0.08 | 0.01 | G | A | 0.73 | 1.40E-08 | 149,821 |
| rs7202877 | 16 | *CTRB1-CTRB2* | 0.11 | 0.02 | T | G | 0.9 | 3.50E-08 | 144,178 |
| rs4812829 | 20 | *HNF4A* | 0.07 | 0.03 | A | G | 0.16 | 5.00E-08 | 77,138 |
| rs7845219 | 8 | *CCNE2; TP53INP1* | 0.08 | 0.02 | T | C | 0.53 | 6.00E-08 | 77,138 |
| rs7961581 | 12 | *TSPAN8* | 0.06 | 0.01 | C | T | 0.27 | 1.80E-07 | 219,582 |
| rs10190052 | 2 | *FAM150B; TMEM18* | 0.07 | 0.02 | C | T | 0.87 | 2.00E-07 | 77,138 |
| rs9472138 | 6 | *TRNAI25* | 0.06 | 0.01 | T | C | 0.25 | 2.00E-07 | 77,138 |
| rs2028299 | 15 | *AP3S2; C15orf38-AP3S2* | 0.04 | 0.02 | C | A | 0.29 | 5.00E-07 | 77,138 |
| rs2820446 | 1 | *RIMKLBP2; ZC3H11B* | 0.05 | 0.01 | C | G | 0.72 | 2.00E-06 | 77,138 |
| rs319598 | 5 | *PCBD2* | 0.05 | 0.01 | C | T | 0.53 | 2.00E-06 | 77,138 |
| rs4273712 | 6 | *YAP1P3; PRELID1P1* | 0.05 | 0.01 | G | A | 0.25 | 3.00E-06 | 77,138 |
| rs12427353 | 12 | *HNF1A* | 0.11 | 0.03 | G | C | 0.79 | 4.00E-06 | 77,138 |
| rs7041847 | 9 | *GLIS3* | 0.05 | 0.02 | A | G | 0.51 | 5.00E-06 | 77,138 |
| rs6937795 | 6 | *SLC35D3; RPL35AP3* | 0.04 | 0.01 | A | C | 0.42 | 7.00E-06 | 77,138 |
| rs1535500 | 6 | *KCNK16; KCNK17* | 0.12 | 0.03 | T | G | 0.59 | 8.00E-06 | 77,138 |
| rs2284219 | 7 | *CRHR2* | 0.05 | 0.01 | G | A | 0.66 | 8.00E-06 | 77,138 |
| rs10788575 | 10 | *RPL11P3; MED6P1* | 0.06 | 0.01 | A | G | 0.17 | 9.00E-06 | 77,138 |
| rs16861329 | 3 | *ST6GAL1* | 0.03 | 0.04 | C | T | 0.85 | 9.00E-06 | 77,138 |

Estimate: log(odds) of T2D per increase in the effect allele, EA: effect allele, OA: other allele, EAF: effect allele frequency, N: sample-size.

**Supplementary Table 3. Association between T2D SNPs and CpGs identified in a meta-EWAS of prevalent T2D, using ALSPAC-ARIES samples (N=1,243). SNP-CpG pairs shown were identified with uncorrected p < 0.05 (Bonferroni p < $1.4x10^{-5}$ or α= 0.05/62 SNPs * 58 CpGs). Associations are ordered by P-value (from smallest to largest).**

| SNP | CpG | SNP gene | Estimate | SE | EA | OA | EAF | P | N |
|---|---|---|---|---|---|---|---|---|---|
| rs4275659 | cg10584271 | ABCB9 | 0.225 | 0.045 | T | C | 0.702 | 5.3E-07 | 1216 |
| rs9936385 | cg14284506 | FTO | 0.158 | 0.041 | C | T | 0.623 | 1.5E-04 | 1242 |
| rs12427353 | cg08945443 | HNF1A | -0.178 | 0.051 | C | G | 0.804 | 5.5E-04 | 1201 |
| rs1359790 | cg20812370 | SPRY2 | 0.161 | 0.047 | A | G | 0.725 | 5.8E-04 | 1216 |
| rs780094 | cg15832662 | GCKR | -0.136 | 0.040 | T | C | 0.586 | 7.5E-04 | 1243 |
| rs2284219 | cg00574958 | CRHR2 | 0.139 | 0.042 | A | G | 0.650 | 8.6E-04 | 1233 |
| rs944801 | cg16192197 | NA | 0.135 | 0.040 | G | C | 0.574 | 8.8E-04 | 1219 |
| rs13266634 | cg04567334 | SLC30A8 | -0.138 | 0.043 | T | C | 0.685 | 1.3E-03 | 1243 |
| rs2284219 | cg00989505 | CRHR2 | 0.130 | 0.042 | A | G | 0.650 | 1.8E-03 | 1233 |
| rs5215 | cg15560632 | KCNJ11 | 0.128 | 0.041 | C | T | 0.630 | 2.1E-03 | 1243 |
| rs3803563 | cg15832662 | PRC1 | -0.164 | 0.053 | A | C | 0.834 | 2.1E-03 | 1236 |
| rs11712037 | cg01577083 | PPARG | -0.189 | 0.062 | G | C | 0.886 | 2.3E-03 | 1233 |
| rs4275659 | cg14003143 | NA | 0.131 | 0.045 | T | C | 0.702 | 3.6E-03 | 1216 |
| rs72999033 | cg14476101 | HAPLN4 | 0.233 | 0.080 | T | C | 0.935 | 3.8E-03 | 1210 |
| rs1496653 | cg11851382 | NA | -0.139 | 0.048 | G | A | 0.782 | 4.0E-03 | 1243 |
| rs17106184 | cg01963618 | FAF1 | -0.192 | 0.067 | A | G | 0.907 | 4.2E-03 | 1236 |
| rs6808574 | cg20116935 | LPP | 0.116 | 0.041 | T | C | 0.623 | 4.6E-03 | 1198 |
| rs35510946 | cg20316538 | IGF2BP2 | 0.122 | 0.043 | A | G | 0.692 | 4.8E-03 | 1229 |
| rs7903146 | cg20456243 | TCF7L2 | -0.122 | 0.044 | T | C | 0.703 | 5.3E-03 | 1243 |
| rs7177055 | cg00162348 | NA | 0.125 | 0.045 | G | A | 0.720 | 5.4E-03 | 1242 |
| rs516946 | cg11376147 | ANK1 | 0.129 | 0.046 | T | C | 0.753 | 5.5E-03 | 1232 |
| rs7961581 | cg00574958 | TSPAN8;LGR5 | 0.125 | 0.045 | C | T | 0.735 | 5.5E-03 | 1235 |
| rs10811660 | cg11851382 | NA | 0.141 | 0.051 | A | G | 0.818 | 5.6E-03 | 1222 |
| rs702634 | cg24704287 | ARL15 | 0.124 | 0.045 | G | A | 0.709 | 5.7E-03 | 1213 |
| rs6937795 | cg15832662 | IL20RA | -0.110 | 0.040 | C | A | 0.521 | 6.5E-03 | 1223 |
| rs516946 | cg16765088 | ANK1 | 0.126 | 0.046 | T | C | 0.753 | 6.6E-03 | 1232 |
| rs72999033 | cg27374726 | HAPLN4 | -0.218 | 0.080 | T | C | 0.935 | 6.7E-03 | 1210 |
| rs4275659 | cg07184465 | NA | 0.122 | 0.045 | T | C | 0.702 | 6.7E-03 | 1216 |
| rs340874 | cg07068382 | PROX1 | 0.110 | 0.041 | T | C | 0.577 | 7.1E-03 | 1242 |
| rs7041847 | cg20154947 | GLIS3 | -0.107 | 0.040 | T | C | 0.522 | 8.1E-03 | 1236 |
| rs10811660 | cg25536676 | NA | -0.134 | 0.051 | A | G | 0.818 | 8.6E-03 | 1222 |
| rs340874 | cg07184465 | PROX1 | 0.107 | 0.041 | T | C | 0.577 | 8.8E-03 | 1242 |
| rs2820446 | cg15832662 | LYPLAL1 | -0.116 | 0.044 | G | C | 0.704 | 8.9E-03 | 1238 |
| rs1974620 | cg19876302 | NA | -0.103 | 0.040 | C | T | 0.528 | 9.3E-03 | 1243 |

**Supplementary Table 3. (*Continued*)**

| SNP | CpG | SNP gene | Estimate | SE | EA | OA | EAF | P | N |
|---|---|---|---|---|---|---|---|---|---|
| rs6937795 | cg18181703 | IL20RA | 0.105 | 0.040 | C | A | 0.521 | 9.5E-03 | 1223 |
| rs11712037 | cg11851382 | PPARG | 0.160 | 0.062 | G | C | 0.886 | 9.5E-03 | 1233 |
| rs77981966 | cg18181703 | THADA | 0.203 | 0.078 | T | C | 0.928 | 9.6E-03 | 1206 |
| rs72999033 | cg08857797 | HAPLN4 | 0.208 | 0.080 | T | C | 0.935 | 9.7E-03 | 1210 |
| rs7903146 | cg01577083 | TCF7L2 | -0.113 | 0.044 | T | C | 0.703 | 9.8E-03 | 1243 |
| rs4275659 | cg24512093 | NA | 0.116 | 0.045 | T | C | 0.702 | 1.0E-02 | 1216 |
| rs7607980 | cg00989505 | COBLL1 | -0.148 | 0.058 | C | T | 0.863 | 1.0E-02 | 1242 |
| rs1535500 | cg12593793 | KCNK16 | 0.100 | 0.039 | T | G | 0.522 | 1.1E-02 | 1229 |
| rs7961581 | cg14476101 | TSPAN8;LGR5 | -0.114 | 0.045 | C | T | 0.735 | 1.1E-02 | 1235 |
| rs10842994 | cg24704287 | KLHDC5 | 0.128 | 0.051 | T | C | 0.804 | 1.2E-02 | 1234 |
| rs7041847 | cg11024682 | GLIS3 | -0.102 | 0.040 | T | C | 0.522 | 1.2E-02 | 1236 |
| rs340874 | cg00144180 | PROX1 | 0.103 | 0.041 | T | C | 0.577 | 1.2E-02 | 1242 |
| rs319598 | cg20116935 | PCBD2 | 0.104 | 0.042 | T | C | 0.582 | 1.2E-02 | 1237 |
| rs5215 | cg14476101 | KCNJ11 | -0.104 | 0.042 | C | T | 0.630 | 1.2E-02 | 1243 |
| rs319598 | cg20316538 | PCBD2 | 0.103 | 0.042 | T | C | 0.582 | 1.4E-02 | 1237 |
| rs5215 | cg08857797 | KCNJ11 | -0.102 | 0.042 | C | T | 0.630 | 1.4E-02 | 1243 |
| rs11257658 | cg06039489 | NA | -0.121 | 0.049 | A | G | 0.776 | 1.4E-02 | 1219 |
| rs2972156 | cg08857797 | NA | -0.102 | 0.042 | C | G | 0.629 | 1.4E-02 | 1221 |
| rs35720761 | cg11252555 | THADA | -0.146 | 0.060 | T | C | 0.871 | 1.5E-02 | 1236 |
| rs7732130 | cg00144180 | ZBED3-AS1 | 0.108 | 0.044 | G | A | 0.697 | 1.5E-02 | 1207 |
| rs5215 | cg00896068 | KCNJ11 | -0.101 | 0.042 | C | T | 0.630 | 1.5E-02 | 1243 |
| rs10510110 | cg01577083 | PLEKHA1 | -0.098 | 0.040 | C | T | 0.526 | 1.5E-02 | 1236 |
| rs702634 | cg00574958 | ARL15 | -0.109 | 0.045 | G | A | 0.709 | 1.5E-02 | 1213 |
| rs2796441 | cg01577083 | TLE1 | -0.100 | 0.042 | A | G | 0.580 | 1.6E-02 | 1187 |
| rs7955901 | cg20316538 | NA | 0.098 | 0.041 | C | T | 0.569 | 1.6E-02 | 1240 |
| rs4275659 | cg27374726 | NA | 0.108 | 0.045 | T | C | 0.702 | 1.6E-02 | 1216 |
| rs12571751 | cg14476101 | ZMIZ1 | 0.098 | 0.041 | G | A | 0.543 | 1.6E-02 | 1233 |
| rs7161785 | cg27374726 | NA | -0.097 | 0.041 | C | G | 0.568 | 1.7E-02 | 1239 |
| rs10811660 | cg00896068 | NA | -0.121 | 0.051 | A | G | 0.818 | 1.7E-02 | 1222 |
| rs1513272 | cg00896068 | NA | -0.096 | 0.040 | C | T | 0.505 | 1.7E-02 | 1231 |
| rs2796441 | cg14284506 | TLE1 | -0.099 | 0.042 | A | G | 0.580 | 1.7E-02 | 1187 |
| rs340874 | cg27115863 | PROX1 | -0.097 | 0.041 | T | C | 0.577 | 1.7E-02 | 1242 |
| rs35720761 | cg08570691 | THADA | -0.142 | 0.060 | T | C | 0.871 | 1.8E-02 | 1236 |
| rs35658696 | cg20154947 | PAM | -0.225 | 0.095 | G | A | 0.952 | 1.8E-02 | 1194 |
| rs1359790 | cg16192197 | SPRY2 | -0.110 | 0.047 | A | G | 0.725 | 1.8E-02 | 1216 |
| rs7955901 | cg20231084 | NA | 0.096 | 0.041 | C | T | 0.569 | 1.8E-02 | 1240 |
| rs340874 | cg00162348 | PROX1 | -0.096 | 0.041 | T | C | 0.577 | 1.9E-02 | 1242 |
| rs17106184 | cg01577083 | FAF1 | 0.158 | 0.067 | A | G | 0.907 | 1.9E-02 | 1236 |
| rs7845219 | cg01963618 | TP53INP1 | -0.093 | 0.039 | T | C | 0.511 | 1.9E-02 | 1229 |

**Supplementary Table 3. (*Continued*)**

| SNP | CpG | SNP gene | Estimate | SE | EA | OA | EAF | P | N |
|---|---|---|---|---|---|---|---|---|---|
| rs17106184 | cg07212837 | FAF1 | 0.157 | 0.067 | A | G | 0.907 | 1.9E-02 | 1236 |
| rs7607980 | cg24795867 | COBLL1 | -0.135 | 0.058 | C | T | 0.863 | 2.0E-02 | 1242 |
| rs9936385 | cg08570691 | FTO | -0.097 | 0.042 | C | T | 0.623 | 2.0E-02 | 1242 |
| rs2972156 | cg06039489 | NA | 0.097 | 0.042 | C | G | 0.629 | 2.0E-02 | 1221 |
| rs516946 | cg25741837 | ANK1 | -0.108 | 0.046 | T | C | 0.753 | 2.0E-02 | 1232 |
| rs1974620 | cg24704287 | NA | -0.092 | 0.040 | C | T | 0.528 | 2.0E-02 | 1243 |
| rs7845219 | cg11024682 | TP53INP1 | 0.092 | 0.039 | T | C | 0.511 | 2.0E-02 | 1229 |
| rs7955901 | cg20154947 | NA | -0.094 | 0.041 | C | T | 0.569 | 2.1E-02 | 1240 |
| rs77981966 | cg16765088 | THADA | 0.182 | 0.079 | T | C | 0.928 | 2.1E-02 | 1206 |
| rs10190052 | cg01963618 | TMEM18 | -0.118 | 0.051 | T | C | 0.813 | 2.1E-02 | 1243 |
| rs35261542 | cg14284506 | CDKAL1 | -0.106 | 0.046 | A | C | 0.740 | 2.1E-02 | 1229 |
| rs340874 | cg07212837 | PROX1 | 0.094 | 0.041 | T | C | 0.577 | 2.1E-02 | 1242 |
| rs1496653 | cg20812370 | NA | 0.110 | 0.048 | G | A | 0.782 | 2.2E-02 | 1243 |
| rs12571751 | cg08570691 | ZMIZ1 | 0.093 | 0.041 | G | A | 0.543 | 2.2E-02 | 1233 |
| rs243020 | cg07212837 | NA | -0.090 | 0.040 | G | A | 0.540 | 2.3E-02 | 1238 |
| rs1974620 | cg14284506 | NA | -0.090 | 0.040 | C | T | 0.528 | 2.3E-02 | 1243 |
| rs9472138 | cg11983038 | VEGFA | -0.101 | 0.045 | T | C | 0.712 | 2.3E-02 | 1243 |
| rs4275659 | cg20231084 | NA | 0.102 | 0.045 | T | C | 0.702 | 2.3E-02 | 1216 |
| rs5215 | cg15832662 | KCNJ11 | -0.093 | 0.042 | C | T | 0.630 | 2.5E-02 | 1243 |
| rs1513272 | cg01577083 | NA | -0.090 | 0.040 | C | T | 0.505 | 2.6E-02 | 1231 |
| rs11712037 | cg14476101 | PPARG | 0.138 | 0.062 | G | C | 0.886 | 2.6E-02 | 1233 |
| rs2028299 | cg06500161 | AP3S2 | 0.098 | 0.044 | C | A | 0.704 | 2.6E-02 | 1235 |
| rs878521 | cg27374726 | YKT6 | -0.106 | 0.048 | A | G | 0.751 | 2.6E-02 | 1202 |
| rs35261542 | cg16575444 | CDKAL1 | 0.102 | 0.046 | A | C | 0.740 | 2.6E-02 | 1229 |
| rs2972156 | cg11851382 | NA | 0.092 | 0.042 | C | G | 0.629 | 2.7E-02 | 1221 |
| rs10276674 | cg20456243 | NA | -0.118 | 0.053 | C | T | 0.816 | 2.7E-02 | 1203 |
| rs7041847 | cg10082515 | GLIS3 | -0.089 | 0.040 | T | C | 0.522 | 2.8E-02 | 1236 |
| rs35720761 | cg04567334 | THADA | -0.131 | 0.060 | T | C | 0.871 | 2.9E-02 | 1236 |
| rs10788575 | cg19693031 | PTEN | -0.117 | 0.054 | A | G | 0.835 | 2.9E-02 | 1236 |
| rs7955901 | cg00574958 | NA | 0.089 | 0.041 | C | T | 0.569 | 2.9E-02 | 1240 |
| rs1552224 | cg00896068 | ARAP1 | 0.118 | 0.054 | C | A | 0.836 | 2.9E-02 | 1243 |
| rs1387153 | cg00896068 | MTNR1B | 0.096 | 0.044 | T | C | 0.707 | 2.9E-02 | 1243 |
| rs7041847 | cg14003143 | GLIS3 | 0.088 | 0.040 | T | C | 0.522 | 2.9E-02 | 1236 |
| rs35510946 | cg11024682 | IGF2BP2 | 0.094 | 0.043 | A | G | 0.692 | 3.0E-02 | 1229 |
| rs12970134 | cg07184465 | MC4R | 0.099 | 0.046 | A | G | 0.739 | 3.0E-02 | 1243 |
| rs944801 | cg20231084 | NA | -0.088 | 0.041 | G | C | 0.574 | 3.0E-02 | 1219 |
| rs13266634 | cg13178597 | SLC30A8 | 0.093 | 0.043 | T | C | 0.685 | 3.1E-02 | 1243 |
| rs6808574 | cg06500161 | LPP | 0.089 | 0.041 | T | C | 0.623 | 3.1E-02 | 1198 |
| rs9472138 | cg06114363 | VEGFA | 0.096 | 0.045 | T | C | 0.712 | 3.1E-02 | 1243 |

### Supplementary Table 3. (*Continued*)

| SNP | CpG | SNP gene | Estimate | SE | EA | OA | EAF | P | N |
|---|---|---|---|---|---|---|---|---|---|
| rs9936385 | cg11024682 | FTO | 0.090 | 0.042 | C | T | 0.623 | 3.1E-02 | 1242 |
| rs944801 | cg00896068 | NA | -0.087 | 0.041 | G | C | 0.574 | 3.1E-02 | 1219 |
| rs35658696 | cg27374726 | PAM | 0.205 | 0.095 | G | A | 0.952 | 3.2E-02 | 1194 |
| rs17106184 | cg11376147 | FAF1 | 0.144 | 0.067 | A | G | 0.907 | 3.2E-02 | 1236 |
| rs10842994 | cg27037013 | KLHDC5 | 0.109 | 0.051 | T | C | 0.804 | 3.2E-02 | 1234 |
| rs12427353 | cg12593793 | HNF1A | 0.110 | 0.052 | C | G | 0.804 | 3.2E-02 | 1201 |
| rs878521 | cg14284506 | YKT6;CAMK2B | 0.102 | 0.048 | A | G | 0.751 | 3.2E-02 | 1202 |
| rs2028299 | cg26270261 | AP3S2 | 0.094 | 0.044 | C | A | 0.704 | 3.2E-02 | 1235 |
| rs4273712 | cg15832662 | C6orf173 | 0.095 | 0.044 | G | A | 0.719 | 3.3E-02 | 1243 |
| rs243020 | cg22680424 | NA | -0.085 | 0.040 | G | A | 0.540 | 3.3E-02 | 1238 |
| rs77981966 | cg11252555 | THADA | -0.168 | 0.079 | T | C | 0.928 | 3.3E-02 | 1206 |
| rs17106184 | cg00989505 | FAF1 | -0.143 | 0.067 | A | G | 0.907 | 3.3E-02 | 1236 |
| rs7961581 | cg18181703 | TSPAN8;LGR5 | 0.096 | 0.045 | C | T | 0.735 | 3.3E-02 | 1235 |
| rs35720761 | cg18181703 | THADA | 0.127 | 0.060 | T | C | 0.871 | 3.3E-02 | 1236 |
| rs4275659 | cg20316538 | NA | 0.096 | 0.045 | T | C | 0.702 | 3.3E-02 | 1216 |
| rs319598 | cg20812370 | PCBD2 | 0.089 | 0.042 | T | C | 0.582 | 3.3E-02 | 1237 |
| rs1387153 | cg25741837 | MTNR1B | 0.093 | 0.044 | T | C | 0.707 | 3.4E-02 | 1243 |
| rs12571751 | cg20316538 | ZMIZ1 | -0.087 | 0.041 | G | A | 0.543 | 3.4E-02 | 1233 |
| rs10190052 | cg25136644 | TMEM18 | 0.108 | 0.051 | T | C | 0.813 | 3.4E-02 | 1243 |
| rs1552224 | cg06114363 | ARAP1,CENTD2 | 0.115 | 0.054 | C | A | 0.836 | 3.5E-02 | 1243 |
| rs9936385 | cg00320980 | FTO | -0.088 | 0.042 | C | T | 0.623 | 3.5E-02 | 1242 |
| rs7202877 | cg07184465 | CTRB1-CTRB2 | -0.138 | 0.066 | G | T | 0.901 | 3.5E-02 | 1243 |
| rs4812829 | cg08570691 | HNF4A | 0.118 | 0.056 | A | G | 0.852 | 3.6E-02 | 1243 |
| rs1552224 | cg07212837 | ARAP1,CENTD2 | 0.114 | 0.054 | C | A | 0.836 | 3.6E-02 | 1243 |
| rs4812829 | cg25136644 | HNF4A | 0.118 | 0.056 | A | G | 0.852 | 3.6E-02 | 1243 |
| rs9936385 | cg11252555 | FTO | -0.087 | 0.042 | C | T | 0.623 | 3.6E-02 | 1242 |
| rs17106184 | cg11983038 | FAF1 | -0.141 | 0.067 | A | G | 0.907 | 3.6E-02 | 1236 |
| rs1552224 | cg27115863 | ARAP1,CENTD2 | 0.113 | 0.054 | C | A | 0.836 | 3.7E-02 | 1243 |
| rs9472138 | cg25536676 | VEGFA | 0.093 | 0.045 | T | C | 0.712 | 3.7E-02 | 1243 |
| rs12571751 | cg09185884 | ZMIZ1 | 0.085 | 0.041 | G | A | 0.543 | 3.7E-02 | 1233 |
| rs944801 | cg11851382 | NA | 0.085 | 0.041 | G | C | 0.574 | 3.7E-02 | 1219 |
| rs702634 | cg07212837 | ARL15 | 0.093 | 0.045 | G | A | 0.709 | 3.7E-02 | 1213 |
| rs11717195 | cg11983038 | ADCY5 | -0.094 | 0.045 | C | T | 0.737 | 3.7E-02 | 1236 |
| rs1513272 | cg14003143 | NA | -0.084 | 0.040 | C | T | 0.505 | 3.7E-02 | 1231 |
| rs7845219 | cg07212837 | TP53INP1 | 0.082 | 0.039 | T | C | 0.511 | 3.8E-02 | 1229 |
| rs243020 | cg26270261 | NA | -0.083 | 0.040 | G | A | 0.540 | 3.8E-02 | 1238 |
| rs10276674 | cg27115863 | NA | -0.111 | 0.053 | C | T | 0.816 | 3.8E-02 | 1203 |

**Supplementary Table 3. (*Continued*)**

| SNP | CpG | SNP gene | Estimate | SE | EA | OA | EAF | P | N |
|---|---|---|---|---|---|---|---|---|---|
| rs77981966 | cg04567334 | THADA | -0.163 | 0.079 | T | C | 0.928 | 3.8E-02 | 1206 |
| rs319598 | cg16192197 | PCBD2 | -0.086 | 0.042 | T | C | 0.582 | 3.8E-02 | 1237 |
| rs1513272 | cg07068382 | NA | 0.084 | 0.040 | C | T | 0.505 | 3.8E-02 | 1231 |
| rs10788575 | cg16575444 | PTEN | -0.111 | 0.054 | A | G | 0.835 | 3.8E-02 | 1236 |
| rs1513272 | cg08570691 | NA | -0.084 | 0.040 | C | T | 0.505 | 3.9E-02 | 1231 |
| rs6937795 | cg26270261 | IL20RA | -0.084 | 0.040 | C | A | 0.521 | 3.9E-02 | 1223 |
| rs2583941 | cg08857797 | RPSAP52 | 0.134 | 0.065 | A | G | 0.897 | 3.9E-02 | 1240 |
| rs7732130 | cg07212837 | ZBED3-AS1 | 0.091 | 0.044 | G | A | 0.697 | 4.0E-02 | 1207 |
| rs319598 | cg08570691 | PCBD2 | 0.085 | 0.042 | T | C | 0.582 | 4.0E-02 | 1237 |
| rs7845219 | cg06039489 | TP53INP1 | -0.081 | 0.039 | T | C | 0.511 | 4.0E-02 | 1229 |
| rs1513272 | cg24686009 | NA | 0.083 | 0.040 | C | T | 0.505 | 4.0E-02 | 1231 |
| rs35720761 | cg27374726 | THADA | -0.122 | 0.060 | T | C | 0.871 | 4.0E-02 | 1236 |
| rs17106184 | cg00144180 | FAF1 | 0.137 | 0.067 | A | G | 0.907 | 4.1E-02 | 1236 |
| rs77981966 | cg08570691 | THADA | -0.161 | 0.079 | T | C | 0.928 | 4.1E-02 | 1206 |
| rs6937795 | cg08945443 | IL20RA | -0.083 | 0.040 | C | A | 0.521 | 4.1E-02 | 1223 |
| rs6937795 | cg00989505 | IL20RA | -0.083 | 0.040 | C | A | 0.521 | 4.1E-02 | 1223 |
| rs340874 | cg20812370 | PROX1 | 0.083 | 0.041 | T | C | 0.577 | 4.2E-02 | 1242 |
| rs1387153 | cg00574958 | MTNR1B | 0.089 | 0.044 | T | C | 0.707 | 4.2E-02 | 1243 |
| rs10842994 | cg16575444 | KLHDC5 | -0.103 | 0.051 | T | C | 0.804 | 4.2E-02 | 1234 |
| rs10842994 | cg25536676 | KLHDC5 | -0.103 | 0.051 | T | C | 0.804 | 4.2E-02 | 1234 |
| rs35720761 | cg22680424 | THADA | -0.121 | 0.060 | T | C | 0.871 | 4.3E-02 | 1236 |
| rs12427353 | cg15560632 | HNF1A | 0.105 | 0.052 | C | G | 0.804 | 4.3E-02 | 1201 |
| rs10788575 | cg07068382 | PTEN | -0.108 | 0.054 | A | G | 0.835 | 4.3E-02 | 1236 |
| rs319598 | cg00162348 | PCBD2 | -0.084 | 0.042 | T | C | 0.582 | 4.3E-02 | 1237 |
| rs11717195 | cg07212837 | ADCY5 | -0.091 | 0.045 | C | T | 0.737 | 4.4E-02 | 1236 |
| rs1974620 | cg14003143 | NA | -0.080 | 0.040 | C | T | 0.528 | 4.4E-02 | 1243 |
| rs1359790 | cg16765088 | SPRY2 | 0.094 | 0.047 | A | G | 0.725 | 4.4E-02 | 1216 |
| rs10190052 | cg22680424 | TMEM18 | 0.102 | 0.051 | T | C | 0.813 | 4.4E-02 | 1243 |
| rs77981966 | cg22680424 | THADA | -0.158 | 0.079 | T | C | 0.928 | 4.4E-02 | 1206 |
| rs1359790 | cg25136644 | SPRY2 | 0.094 | 0.047 | A | G | 0.725 | 4.5E-02 | 1216 |
| rs1387153 | cg16192197 | MTNR1B | 0.088 | 0.044 | T | C | 0.707 | 4.5E-02 | 1243 |
| rs2583941 | cg00896068 | RPSAP52 | -0.131 | 0.065 | A | G | 0.897 | 4.5E-02 | 1240 |
| rs11717195 | cg06114363 | ADCY5 | 0.091 | 0.045 | C | T | 0.737 | 4.5E-02 | 1236 |
| rs10811660 | cg20231084 | NA | -0.102 | 0.051 | A | G | 0.818 | 4.5E-02 | 1222 |
| rs1513272 | cg27037013 | NA | -0.081 | 0.040 | C | T | 0.505 | 4.5E-02 | 1231 |
| rs6937795 | cg20116935 | IL20RA | -0.081 | 0.040 | C | A | 0.521 | 4.5E-02 | 1223 |

**Supplementary Table 3. (*Continued*)**

| SNP | CpG | SNP gene | Estimate | SE | EA | OA | EAF | P | N |
|---|---|---|---|---|---|---|---|---|---|
| rs1552224 | cg27374726 | ARAP1,CENTD2 | 0.109 | 0.054 | C | A | 0.836 | 4.5E-02 | 1243 |
| rs35261542 | cg22680424 | CDKAL1 | -0.092 | 0.046 | A | C | 0.740 | 4.5E-02 | 1229 |
| rs4275659 | cg01963618 | NA | 0.090 | 0.045 | T | C | 0.702 | 4.5E-02 | 1216 |
| rs1496653 | cg11376147 | NA | -0.096 | 0.048 | G | A | 0.782 | 4.6E-02 | 1243 |
| rs9936385 | cg26766064 | FTO | -0.083 | 0.042 | C | T | 0.623 | 4.6E-02 | 1242 |
| rs1387153 | cg01963618 | MTNR1B | -0.087 | 0.044 | T | C | 0.707 | 4.6E-02 | 1243 |
| rs7607980 | cg19693031 | COBLL1 | -0.115 | 0.058 | C | T | 0.863 | 4.6E-02 | 1242 |
| rs13266634 | cg25536676 | SLC30A8 | 0.086 | 0.043 | T | C | 0.685 | 4.7E-02 | 1243 |
| rs41278853 | cg11851382 | MTMR3 | -0.131 | 0.066 | G | A | 0.895 | 4.7E-02 | 1242 |
| rs2284219 | cg01577083 | CRHR2 | 0.083 | 0.042 | A | G | 0.650 | 4.7E-02 | 1233 |
| rs1535500 | cg07068382 | KCNK16 | 0.078 | 0.039 | T | G | 0.522 | 4.8E-02 | 1229 |
| rs2284219 | cg11983038 | CRHR2 | -0.083 | 0.042 | A | G | 0.650 | 4.8E-02 | 1233 |
| rs10190052 | cg06500161 | TMEM18 | 0.101 | 0.051 | T | C | 0.813 | 4.8E-02 | 1243 |
| rs1974620 | cg25136644 | NA | -0.078 | 0.040 | C | T | 0.528 | 4.9E-02 | 1243 |
| rs4275659 | cg22680424 | NA | 0.089 | 0.045 | T | C | 0.702 | 4.9E-02 | 1216 |
| rs12571751 | cg20116935 | ZMIZ1 | 0.080 | 0.041 | G | A | 0.543 | 4.9E-02 | 1233 |
| rs77981966 | cg00144180 | THADA | -0.155 | 0.079 | T | C | 0.928 | 4.9E-02 | 1206 |
| rs17106184 | cg22680424 | FAF1 | 0.132 | 0.067 | A | G | 0.907 | 4.9E-02 | 1236 |
| rs780094 | cg20812370 | GCKR | -0.079 | 0.040 | T | C | 0.586 | 5.0E-02 | 1243 |
| rs702634 | cg01577083 | ARL15 | 0.088 | 0.045 | G | A | 0.709 | 5.0E-02 | 1213 |
| rs35720761 | cg20231084 | THADA | -0.117 | 0.060 | T | C | 0.871 | 5.0E-02 | 1236 |

Estimate: additive effect of the genotype on a unit change in inverse-normal transformed residuals of DNA methylation, EA: effect allele, OA: other allele, EAF: effect allele frequency, N: sample-size.

**Supplementary Table 4. Methylation quantitative trait loci (mQTL) associated with a subset of the meta-EWAS of T2D CpGs. mQTL were retrieved from the Genetics of DNAm consortium (GoDMC). In total, 39 mQTL proxying 30 of the 58 meta-EWAS CpGs, were used as instruments in the reverse two-sample MR analysis (DNAm → T2D). Associations are ordered by P-value (from the smallest to the largest).**

| SNP | CpG | CpG gene | Cis/Trans | Estimate | SE | EA | OA | EAF | P | N |
|---|---|---|---|---|---|---|---|---|---|---|
| rs11693641 | cg00144180 | *HDAC4* | Cis | -0.150 | 0.010 | A | C | 0.5 | 3.5E-202 | 23360 |
| rs113786621 | cg00896068 | *Open sea* | Cis | -0.340 | 0.020 | T | C | 0.08 | 3.5E-202 | 25956 |
| rs56261297 | cg07212837 | *Open sea* | Cis | -0.340 | 0.010 | T | C | 0.41 | 3.5E-202 | 27738 |
| rs115738369 | cg10584271 | *ITIH1* | Cis | -1.740 | 0.030 | T | C | 0.02 | 3.5E-202 | 22070 |
| rs11652574 | cg11024682 | *SREBF1* | Cis | 1.100 | 0.030 | A | G | 0.04 | 3.5E-202 | 19085 |
| rs11584621 | cg12593793 | *Open sea* | Cis | -0.060 | 0.010 | A | T | 0.21 | 3.5E-202 | 25084 |
| rs347903 | cg14476101 | *PHGDH* | Cis | 0.230 | 0.010 | T | C | 0.67 | 3.5E-202 | 24554 |
| rs750129 | cg20231084 | *Open sea* | Cis | 0.140 | 0.010 | A | G | 0.47 | 3.5E-202 | 23944 |
| rs55760516 | cg20456243 | *SPEG* | Cis | -0.120 | 0.010 | A | G | 0.67 | 3.5E-202 | 27242 |
| rs9309801 | cg24512093 | *ROBO1* | Cis | -0.110 | 0.010 | T | C | 0.34 | 3.5E-202 | 27235 |
| rs9831014 | cg24512093 | *ROBO1* | Cis | 0.120 | 0.010 | C | G | 0.42 | 3.5E-202 | 24994 |
| rs6681644 | cg25536676 | *DHCR24* | Cis | 0.260 | 0.010 | C | G | 0.42 | 3.5E-202 | 27714 |
| rs13051329 | cg27037013 | *Open sea* | Cis | 0.170 | 0.010 | T | C | 0.15 | 3.5E-202 | 26837 |
| rs6000773 | cg27115863 | *Open sea* | Cis | -0.120 | 0.010 | C | G | 0.75 | 3.5E-202 | 24389 |
| rs9487736 | cg16192197 | *Open sea* | Cis | 0.360 | 0.010 | A | G | 0.14 | 6.3E-188 | 27726 |
| rs608358 | cg14476101 | *PHGDH* | Cis | -0.280 | 0.010 | A | C | 0.28 | 5.8E-180 | 25566 |
| rs1525502 | cg10082515 | *Open sea* | Cis | -0.250 | 0.010 | T | C | 0.41 | 1.1E-172 | 24607 |
| rs7496161 | cg16765088 | *Open sea* | Cis | -0.320 | 0.010 | A | G | 0.11 | 4.7E-122 | 25984 |
| rs1107095 | cg01577083 | *Open sea* | Cis | -0.200 | 0.010 | T | C | 0.51 | 7.5E-109 | 23764 |
| rs7602568 | cg27115863 | *Open sea* | Trans | 0.200 | 0.010 | T | C | 0.22 | 1.0E-85 | 27625 |
| rs4383852 | cg18181703 | *SOCS3* | Trans | -0.130 | 0.010 | A | G | 0.52 | 2.3E-56 | 27746 |
| rs6596785 | cg01963618 | *LOC285768* | Cis | 0.220 | 0.010 | A | G | 0.89 | 2.2E-50 | 25226 |
| rs1047891 | cg08857797 | *VPS25* | Trans | -0.140 | 0.010 | A | C | 0.32 | 8.8E-47 | 24138 |
| rs9525281 | cg00896068 | *Open sea* | Cis | 0.140 | 0.010 | C | G | 0.76 | 3.2E-30 | 18956 |
| rs174551 | cg25536676 | *DHCR24* | Trans | 0.110 | 0.010 | T | C | 0.66 | 5.1E-30 | 24653 |
| rs10421294 | cg11252555 | *RPL13AP5* | Cis | -0.140 | 0.010 | A | G | 0.1 | 2.8E-20 | 25481 |
| rs540908 | cg13178597 | *RGS17* | Cis | -0.100 | 0.010 | A | G | 0.82 | 6.4E-18 | 26442 |
| rs2848634 | cg11376147 | *SLC43A1* | Cis | 0.090 | 0.010 | A | G | 0.75 | 7.9E-18 | 27749 |
| rs71380866 | cg09185884 | *KCTD2* | Cis | 0.210 | 0.030 | C | G | 0.97 | 8.7E-15 | 25599 |
| rs62250760 | cg10584271 | *ITIH1* | Cis | 0.050 | 0.010 | T | C | 0.35 | 9.1E-08 | 25857 |
| rs79365581 | cg25536676 | *DHCR24* | Cis | 0.350 | 0.070 | T | C | 0.98 | 7.8E-07 | 5834 |
| rs1872614 | cg00144180 | *HDAC4* | Cis | -0.050 | 0.010 | A | T | 0.58 | 2.2E-06 | 16512 |
| rs6081870 | cg06039489 | *C20orf26* | Cis | -0.141 | 0.010 | A | G | 0.2685 | 7.3E-44 | 24388 |
| rs220182 | cg06500161 | *ABCG1* | Cis | 0.061 | 0.009 | T | C | 0.5528 | 3.5E-202 | 24474 |
| rs1500138 | cg07184465 | *SPZ1* | Cis | -0.147 | 0.009 | T | C | 0.3522 | 1.8E-58 | 25936 |

**Supplementary Table 4. (*Continued*)**

| SNP | CpG | CpG gene | Cis/Trans | Estimate | SE | EA | OA | EAF | P | N |
|---|---|---|---|---|---|---|---|---|---|---|
| rs7535757 | cg11851382 | *PPAP2B* | Cis | -0.077 | 0.009 | A | G | 0.4988 | 3.5E-202 | 26658 |
| rs6657798 | cg19693031 | *TXNIP* | Trans | -0.459 | 0.010 | C | G | 0.8024 | 3.5E-202 | 27212 |
| rs6732515 | cg25741837 | *SMYD5* | Cis | 0.542 | 0.032 | A | C | 0.9767 | 1.1E-64 | 22690 |
| rs62148128 | cg25741837 | *SMYD5* | Cis | 0.715 | 0.021 | A | G | 0.0613 | 3.5E-202 | 20444 |

Estimate: additive effect of the genotype on a unit change in inverse-normal transformed residuals of DNA methylation as reported by the GoDMC consortium, EA: effect allele, OA: other allele, EAF: effect allele frequency, N: sample-size.

**Supplementary Table 5. Association of mQTL SNPs tagging meta-EWAS CpGs with prevalent T2D using GWAS summary data. GWAS of T2D data was extracted from Mahajan *et al.* 2014 and Wood *et al.* 2016. In total, T2D associations were retrieved for 39 mQTL SNPs proxying 30 meta-EWAS CpGs. Data is ordered by P-value (from smallest to largest).**

| SNP | Exposure | Outcome | T2D GWAS | EA | OA | EAF | Estimate | SE | P | N |
|---|---|---|---|---|---|---|---|---|---|---|
| rs174551*† | cg25536676 | T2D | Mahajan et al. 2014 | T | C | NA | 0.039 | 0.012 | 0.011 | 104377 |
| rs7496161† | cg16765088 | T2D | Wood et al. 2016 | A | G | 0.10 | -0.174 | 0.085 | 0.034 | 116171 |
| rs1872614† | cg00144180 | T2D | Wood et al. 2016 | A | T | 0.58 | -0.095 | 0.036 | 0.037 | 104810 |
| rs1047891† | cg08857797 | T2D | Wood et al. 2016 | A | C | 0.32 | -0.073 | 0.038 | 0.042 | 117775 |
| rs6681644 | cg25536676 | T2D | Wood et al. 2016 | C | G | 0.42 | 0.095 | 0.033 | 0.054 | 117094 |
| rs347903* | cg14476101 | T2D | Mahajan et al. 2014 | T | C | NA | -0.030 | 0.015 | 0.057 | 107656 |
| rs1500138 | cg07184465 | T2D | Wood et al. 2016 | T | C | 0.35 | -0.062 | 0.035 | 0.072 | 117682 |
| rs56261297* | cg07212837 | T2D | Mahajan et al. 2014 | T | C | NA | -0.030 | 0.012 | 0.072 | 96027 |
| rs1525502 | cg10082515 | T2D | Wood et al. 2016 | T | C | 0.39 | 0.095 | 0.034 | 0.098 | 115383 |
| rs7535757 | cg11851382 | T2D | Wood et al. 2016 | A | G | 0.49 | 0.095 | 0.033 | 0.100 | 115683 |
| rs10421294* | cg11252555 | T2D | Mahajan et al. 2014 | A | G | NA | 0.039 | 0.025 | 0.110 | 95397 |
| rs1525502 | cg10082515 | T2D | Mahajan et al. 2014 | T | C | NA | 0.020 | 0.015 | 0.120 | 105669 |
| rs1047891* | cg08857797 | T2D | Mahajan et al. 2014 | A | C | NA | -0.039 | 0.025 | 0.150 | 24243 |
| rs9525281 | cg00896068 | T2D | Wood et al. 2016 | C | G | 0.75 | 0.062 | 0.046 | 0.160 | 109280 |
| rs7535757 | cg11851382 | T2D | Mahajan et al. 2014 | A | G | NA | 0.020 | 0.013 | 0.170 | 102682 |
| rs62250760* | cg10584271 | T2D | Mahajan et al. 2014 | T | C | NA | -0.020 | 0.015 | 0.170 | 106223 |
| rs11693641 | cg00144180 | T2D | Wood et al. 2016 | A | C | 0.49 | -0.041 | 0.032 | 0.180 | 117061 |
| rs79365581 | cg25536676 | T2D | Wood et al. 2016 | T | C | 0.99 | 0.734 | 0.540 | 0.180 | 117420 |
| rs11693641* | cg00144180 | T2D | Mahajan et al. 2014 | A | C | NA | -0.020 | 0.013 | 0.180 | 102563 |
| rs2848634 | cg11376147 | T2D | Mahajan et al. 2014 | A | G | NA | -0.020 | 0.015 | 0.210 | 100365 |
| rs56261297 | cg07212837 | T2D | Wood et al. 2016 | T | C | 0.41 | 0.000 | 0.033 | 0.270 | 117775 |
| rs6000773 | cg27115863 | T2D | Wood et al. 2016 | C | G | 0.74 | 0.000 | 0.042 | 0.280 | 116130 |
| rs113786621 | cg00896068 | T2D | Wood et al. 2016 | T | C | 0.08 | 0.095 | 0.120 | 0.300 | 116849 |
| rs6081870 | cg06039489 | T2D | Wood et al. 2016 | A | G | 0.27 | 0.000 | 0.042 | 0.300 | 113116 |
| rs55760516* | cg20456243 | T2D | Mahajan et al. 2014 | A | G | NA | -0.010 | 0.013 | 0.300 | 104314 |
| rs6732515* | cg25741837 | T2D | Mahajan et al. 2014 | A | C | NA | 0.058 | 0.067 | 0.350 | 63390 |
| rs6732515 | cg25741837 | T2D | Wood et al. 2016 | A | C | 0.98 | 0.288 | 0.310 | 0.360 | 116210 |
| rs10421294 | cg11252555 | T2D | Wood et al. 2016 | A | G | 0.09 | 0.095 | 0.099 | 0.370 | 117526 |
| rs13051329 | cg27037013 | T2D | Wood et al. 2016 | T | C | 0.15 | -0.051 | 0.064 | 0.380 | 117504 |
| rs174551 | cg25536676 | T2D | Wood et al. 2016 | T | C | 0.65 | 0.000 | 0.036 | 0.400 | 117520 |
| rs55760516 | cg20456243 | T2D | Wood et al. 2016 | A | G | 0.68 | 0.000 | 0.037 | 0.400 | 117775 |
| rs1107095 | cg01577083 | T2D | Wood et al. 2016 | T | C | 0.51 | 0.000 | 0.033 | 0.410 | 110804 |
| rs62250760 | cg10584271 | T2D | Wood et al. 2016 | T | C | 0.35 | -0.030 | 0.036 | 0.420 | 116430 |
| rs11652574 | cg11024682 | T2D | Wood et al. 2016 | A | G | 0.03 | -0.198 | 0.240 | 0.420 | 117291 |

**Supplementary Table 5. (*Continued*)**

| SNP | Exposure | Outcome | T2D GWAS | EA | OA | EAF | Estimate | SE | P | N |
|---|---|---|---|---|---|---|---|---|---|---|
| rs347903 | cg14476101 | T2D | Wood et al. 2016 | T | C | 0.65 | 0.000 | 0.037 | 0.420 | 111060 |
| rs9309801 | cg24512093 | T2D | Wood et al. 2016 | T | C | 0.33 | 0.000 | 0.037 | 0.420 | 116463 |
| rs4383852 | cg18181703 | T2D | Wood et al. 2016 | A | G | 0.52 | 0.030 | 0.032 | 0.420 | 117641 |
| rs7602568 | cg27115863 | T2D | Wood et al. 2016 | T | C | 0.23 | -0.030 | 0.046 | 0.450 | 117775 |
| rs540908 | cg13178597 | T2D | Wood et al. 2016 | A | G | 0.81 | 0.000 | 0.055 | 0.470 | 116958 |
| rs6081870 | cg06039489 | T2D | Mahajan et al. 2014 | A | G | NA | 0.010 | 0.015 | 0.500 | 106232 |
| rs7496161 | cg16765088 | T2D | Mahajan et al. 2014 | A | G | NA | 0.010 | 0.018 | 0.510 | 98254 |
| rs6596785 | cg01963618 | T2D | Mahajan et al. 2014 | A | G | NA | 0.010 | 0.018 | 0.540 | 104567 |
| rs9831014 | cg24512093 | T2D | Wood et al. 2016 | C | G | 0.42 | -0.020 | 0.034 | 0.580 | 115128 |
| rs220182 | cg06500161 | T2D | Mahajan et al. 2014 | T | C | NA | -0.010 | 0.015 | 0.620 | 91284 |
| rs9487736 | cg16192197 | T2D | Mahajan et al. 2014 | A | G | NA | -0.010 | 0.015 | 0.630 | 100236 |
| rs1500138 | cg07184465 | T2D | Mahajan et al. 2014 | T | C | NA | 0.010 | 0.015 | 0.640 | 104490 |
| rs2848634 | cg11376147 | T2D | Wood et al. 2016 | A | G | 0.75 | 0.000 | 0.042 | 0.680 | 117587 |
| rs6657798 | cg19693031 | T2D | Wood et al. 2016 | C | G | 0.82 | 0.000 | 0.056 | 0.710 | 116594 |
| rs62148128 | cg25741837 | T2D | Wood et al. 2016 | A | G | 0.06 | 0.095 | 0.170 | 0.750 | 116700 |
| rs750129 | cg20231084 | T2D | Mahajan et al. 2014 | A | G | NA | 0.000 | 0.013 | 0.770 | 106230 |
| rs608358 | cg14476101 | T2D | Wood et al. 2016 | A | C | 0.28 | 0.000 | 0.041 | 0.780 | 115429 |
| rs750129 | cg20231084 | T2D | Wood et al. 2016 | A | G | 0.47 | -0.010 | 0.036 | 0.800 | 96646 |
| rs13051329 | cg27037013 | T2D | Mahajan et al. 2014 | T | C | NA | 0.000 | 0.018 | 0.800 | 104328 |
| rs4383852* | cg18181703 | T2D | Mahajan et al. 2014 | A | G | NA | 0.000 | 0.013 | 0.810 | 98320 |
| rs9309801 | cg24512093 | T2D | Mahajan et al. 2014 | T | C | NA | 0.000 | 0.013 | 0.810 | 104489 |
| rs11584621 | cg12593793 | T2D | Wood et al. 2016 | A | T | 0.21 | -0.010 | 0.051 | 0.860 | 113096 |
| rs7602568 | cg27115863 | T2D | Mahajan et al. 2014 | T | C | NA | 0.000 | 0.015 | 0.910 | 110135 |
| rs71380866 | cg09185884 | T2D | Wood et al. 2016 | C | G | 0.97 | 0.030 | 0.300 | 0.920 | 117447 |
| rs115738369 | cg10584271 | T2D | Wood et al. 2016 | T | C | 0.02 | -0.030 | 0.520 | 0.950 | 117430 |
| rs6596785 | cg01963618 | T2D | Wood et al. 2016 | A | G | 0.90 | 0.010 | 0.097 | 0.950 | 115281 |
| rs608358 | cg14476101 | T2D | Mahajan et al. 2014 | A | C | NA | 0.000 | 0.015 | 0.970 | 101903 |
| rs220182 | cg06500161 | T2D | Wood et al. 2016 | T | C | 0.54 | 0.000 | 0.033 | 1.000 | 114976 |

Estimate: effect of the genotype on the log(odds) of prevalent T2D as reported in two previous GWAS [9, 10], EA: effect allele of the outcome, OA: other allele of the outcome, EAF: effect allele frequency of the outcome, N: sample-size of the outcome. *mQTL SNPs tagged by an alternative SNP in high LD ($r^2$ >= 0.6) that was found in the outcome dataset (GWAS of T2D) in MR-Base. Genetic proxies for the mQTL SNPs rs347903, rs10421294, rs1047891, rs174551, rs11693641, rs4383852, rs55760516, rs56261297, rs6732515 and rs62250760, were tagged by the SNPs rs838990, rs4802613, rs715, rs1535, rs1399629, rs2884013, rs1050816, rs7459603, rs6727270 and rs13314396, respectively, using data from the 1,000 genomes in Europeans. †SNPs associated with T2D with borderline GWAS significance at $P < 0.05$.

**Supplementary Table 6. Results of the reverse two-sample MR (DNAm → T2D) using additional MR methods at the CpG cg25536676 (*DHCR24*). Three mQTL were identified as genetic proxies for this CpG in the GoDMC data set.**

| Outcome | T2D GWAS | Method | # SNPs | OR (95%CI) | P | Het | P-Het |
|---|---|---|---|---|---|---|---|
| T2D | Mahajan et al. 2014 | Wald ratio | 1 | 1.43(1.15,1.78) | 0.001 | | |
| T2D | Wood et al. 2016 | IVW | 3 | 1.39(1.08,1.79) | 0.011 | 2.41 | 0.30 |
| T2D | Wood et al. 2016 | MR Egger | 3 | 1.95(0.99,3.85) | 0.305 | 1.15 | 0.28 |
| T2D | Wood et al. 2016 | Weighted median | 3 | 1.38(1.1,1.73) | 0.006 | | |
| T2D | Wood et al. 2016 | Weighted mode | 3 | 1.39(1.1,1.75) | 0.107 | | |

IVW: inverse variance-weighted regression. # SNPs: number of single nucleotide polymorphisms used to proxy the CpG cg25536676 (*DHCR24*) and included in the MR analysis. Het: heterogeneity value calculated using the Cochran's Q and Rucker's estimates for the IVW and MR Egger regressions, respectively. P-Het: P value of the heterogeneity test. Effect estimates are expressed in odds ratios of T2D per increase in inverse normal-transformed residuals of DNA methylation at cg25536676.

**Supplementary Table 7. Comparison of association estimates between the observational and the causal analysis (bidirectional two-sample MR) for 58 CpGs previously reported in a meta-EWAS of prevalent T2D ($P < 1.3x10^{-5}$). Likely causal direction of association was established based on the smallest P-value, and on the directional consistency found between the causal and the observational effect estimates for each direction of the 2SMR. *Inconclusive – single direction* if MR data was only available in one direction and $P > 0.1$; *inconclusive-bidirectional* if MR data was available in two directions, but $P > 0.1$ in both cases.**

| | | | **Meta-EWAS of T2D (N= 3,248)** | | **Forward 2SMR (T2D → DNAm)** | | **Reverse 2SMR (DNAm → T2D)** | | **Likely causal direction** |
|---|---|---|---|---|---|---|---|---|---|
| CpG | Chr | Gene | Estimate (95%CI) | P | Estimate (95%CI) | P | Estimate (95%CI) | P | |
| cg06114363 | 1 | *ZNF683* | -0.01(-0.015,-0.006) | 1.37E-06 | -0.017(-0.13,0.1) | 0.778 | NA | NA | Inconclusive - single direction |
| cg11851382** | 1 | *PPAP2B* | -0.008(-0.011,-0.004) | 6.42E-06 | 0.031(-0.09,0.15) | 0.603 | -1.24(-2.09,-0.4) | 0.004 | DNAm to T2D |
| cg12593793 | 1 | *Open sea* | -0.008(-0.011,-0.004) | 2.90E-06 | 0.039(-0.08,0.16) | 0.519 | 0.17(-1.5,1.83) | 0.844 | T2D to DNAm |
| cg14476101 | 1 | *PHGDH* | -0.015(-0.021,-0.008) | 9.46E-06 | -0.008(-0.14,0.12) | 0.901 | -0.05(-0.18,0.07) | 0.398 | Inconclusive - bidirectional |
| cg19693031† | 1 | *TXNIP* | -0.019(-0.024,-0.014) | 8.75E-14 | -0.035(-0.15,0.08) | 0.555 | 0.00(-0.24,0.24) | 1.000 | Inconclusive - bidirectional |
| cg20812370* | 1 | *PBX1* | -0.007(-0.009,-0.004) | 7.40E-07 | -0.184(-0.3,-0.07) | 0.002 | NA | NA | T2D to DNAm |
| cg25536676** | 1 | *DHCR24* | -0.008(-0.011,-0.004) | 5.39E-06 | 0.042(-0.08,0.16) | 0.490 | 0.36(0.14,0.58) | 0.001 | DNAm to T2D |
| cg00144180†‡ | 2 | *HDAC4* | 0.012(0.008,0.017) | 5.64E-08 | 0.071(-0.05,0.19) | 0.256 | 0.13(-0.03,0.3) | 0.115 | DNAm to T2D |
| cg20316538 | 2 | *RUFY4* | -0.005(-0.007,-0.003) | 6.11E-06 | -0.013(-0.14,0.11) | 0.841 | NA | NA | Inconclusive - single direction |
| cg20456243* | 2 | *SPEG* | -0.007(-0.011,-0.004) | 9.99E-06 | -0.066(-0.19,0.06) | 0.304 | 0.08(-0.12,0.29) | 0.429 | T2D to DNAm |
| cg25741837 | 2 | *SMYD5* | 0.009(0.005,0.013) | 4.76E-06 | 0.043(-0.07,0.16) | 0.474 | 0.11(-0.14,0.35) | 0.385 | DNAm to T2D |
| cg10584271 | 3 | *ITIH1* | -0.014(-0.019,-0.009) | 1.73E-07 | -0.068(-0.19,0.06) | 0.279 | -0.4(-0.98,0.19) | 0.187 | DNAm to T2D |
| cg20116935 | 3 | *SEMA3B* | -0.006(-0.009,-0.003) | 8.89E-06 | -0.047(-0.17,0.07) | 0.430 | NA | NA | Inconclusive - single direction |
| cg24512093 | 3 | *ROBO1* | -0.01(-0.013,-0.006) | 7.16E-07 | -0.036(-0.15,0.08) | 0.546 | 0.00(-0.23,0.23) | 1.000 | T2D to DNAm |
| cg07184465 | 5 | *SPZ1* | -0.007(-0.01,-0.004) | 7.18E-06 | -0.051(-0.17,0.07) | 0.403 | 0.42(-0.05,0.89) | 0.077 | DNAm to T2D |
| cg01963618 | 6 | *LOC285768* | -0.008(-0.011,-0.004) | 1.55E-06 | -0.002(-0.12,0.12) | 0.970 | 0.05(-0.11,0.2) | 0.572 | T2D to DNAm |
| cg07068382 | 6 | *MTCH1* | 0.01(0.006,0.015) | 9.46E-06 | 0.039(-0.08,0.16) | 0.529 | NA | NA | Inconclusive - single direction |
| cg13178597 | 6 | *RGS17* | -0.01(-0.015,-0.006) | 8.57E-06 | -0.035(-0.15,0.08) | 0.560 | 0.00(-1.08,1.08) | 1.000 | T2D to DNAm |
| cg16192197 | 6 | *Open sea* | 0.01(0.005,0.014) | 3.71E-06 | -0.017(-0.14,0.11) | 0.784 | -0.03(-0.11,0.05) | 0.512 | DNAm to T2D |
| cg10082515** | 7 | *Open sea* | -0.013(-0.019,-0.008) | 7.46E-06 | -0.046(-0.16,0.07) | 0.443 | -0.38(-0.65,-0.11) | 0.005 | DNAm to T2D |
| cg15560632 | 7 | *LRCH4* | -0.001(-0.001,-0.001) | 3.83E-06 | -0.038(-0.16,0.08) | 0.530 | NA | NA | Inconclusive - single direction |

### Supplementary Table 7. (*Continued*)

| CpG | Chr | Gene | Meta-EWAS of T2D (N= 3,248) Estimate (95%CI) | P | Forward 2SMR (T2D → DNAm) Estimate (95%CI) | P | Reverse 2SMR (DNAm → T2D) Estimate (95%CI) | P | Likely causal direction |
|---|---|---|---|---|---|---|---|---|---|
| cg25136644 | 7 | *ATG9B* | -0.007(-0.01,-0.004) | 7.27E-07 | -0.09(-0.21,0.03) | 0.132 | NA | NA | Inconclusive - single direction |
| cg07212837** | 8 | *Open sea* | 0.006(0.004,0.009) | 3.28E-06 | -0.069(-0.2,0.06) | 0.304 | 0.09(0.02,0.16) | 0.018 | DNAm to T2D |
| cg20154947 | 8 | *PLEC1* | -0.002(-0.003,-0.001) | 4.34E-06 | 0.005(-0.11,0.12) | 0.938 | NA | NA | Inconclusive - single direction |
| cg00320980 | 10 | *Open sea* | -0.009(-0.013,-0.005) | 7.97E-06 | -0.045(-0.16,0.07) | 0.449 | NA | NA | Inconclusive - single direction |
| cg04567334 | 10 | *CDH23* | -0.006(-0.008,-0.004) | 1.67E-07 | 0.035(-0.09,0.16) | 0.567 | NA | NA | Inconclusive - single direction |
| cg08945443 | 10 | *ZMYND17* | 0.01(0.006,0.015) | 2.64E-06 | 0.005(-0.12,0.13) | 0.939 | NA | NA | Inconclusive - single direction |
| cg19876302 | 10 | *Open sea* | -0.008(-0.011,-0.005) | 2.22E-06 | 0.045(-0.07,0.16) | 0.450 | NA | NA | Inconclusive - single direction |
| cg27374726 | 10 | *Open sea* | -0.009(-0.012,-0.005) | 2.32E-06 | -0.079(-0.2,0.04) | 0.187 | NA | NA | Inconclusive - single direction |
| cg00574958† | 11 | *CPT1A* | -0.007(-0.01,-0.005) | 1.20E-08 | 0.008(-0.13,0.14) | 0.915 | NA | NA | Inconclusive - single direction |
| cg11376147 | 11 | *SLC43A1* | -0.006(-0.008,-0.003) | 5.43E-06 | -0.045(-0.16,0.07) | 0.454 | -0.22(-0.55,0.11) | 0.187 | DNAm to T2D |
| cg15832662 | 11 | *RTN3* | -0.009(-0.013,-0.005) | 8.45E-06 | -0.005(-0.14,0.13) | 0.945 | NA | NA | Inconclusive - single direction |
| cg20231084 | 11 | *Open sea* | -0.006(-0.009,-0.003) | 8.36E-06 | 0.01(-0.11,0.13) | 0.871 | -0.07(-0.58,0.43) | 0.780 | DNAm to T2D |
| cg22680424 | 11 | *HCCA2* | 0.008(0.005,0.011) | 2.16E-06 | -0.099(-0.22,0.02) | 0.109 | NA | NA | Inconclusive - single direction |
| cg24686009* | 12 | *RAP1B* | -0.002(-0.003,-0.001) | 1.19E-06 | 0.004(-0.11,0.12) | 0.942 | NA | NA | Inconclusive - single direction |
| cg24795867 | 12 | *WNT5B* | -0.006(-0.009,-0.004) | 2.47E-06 | 0.045(-0.07,0.16) | 0.450 | NA | NA | Inconclusive - single direction |
| cg26270261 | 12 | *KRT4* | -0.006(-0.009,-0.004) | 5.68E-07 | 0.05(-0.07,0.17) | 0.418 | NA | NA | Inconclusive - single direction |
| cg00896068 | 13 | *Open sea* | -0.008(-0.011,-0.004) | 7.58E-06 | -0.077(-0.2,0.05) | 0.233 | 0.11(-0.6,0.81) | 0.767 | Inconclusive - bidirectional |
| cg11983038 | 13 | *Open sea* | -0.017(-0.023,-0.01) | 7.23E-07 | 0.039(-0.09,0.16) | 0.539 | NA | NA | Inconclusive - single direction |
| cg00989505 | 14 | *MIR299* | -0.004(-0.006,-0.002) | 9.33E-06 | -0.034(-0.16,0.09) | 0.601 | NA | NA | Inconclusive - single direction |
| cg16765088**† | 15 | *SYNM* | -0.011(-0.014,-0.007) | 5.50E-10 | -0.079(-0.2,0.04) | 0.188 | 0.54(0.02,1.07) | 0.040 | DNAm to T2D |
| cg00162348 | 16 | *RNF40* | -0.002(-0.003,-0.001) | 6.64E-06 | 0.018(-0.1,0.14) | 0.766 | NA | NA | Inconclusive - single direction |
| cg01577083* | 16 | *Open sea* | -0.011(-0.016,-0.006) | 7.93E-06 | -0.151(-0.28,-0.02) | 0.023 | 0.00(-0.32,0.32) | 1.000 | T2D to DNAm |
| cg16575444 | 16 | *CX3CL1* | -0.006(-0.008,-0.004) | 6.83E-07 | 0.01(-0.11,0.13) | 0.862 | NA | NA | Inconclusive - single direction |
| cg08857797 | 17 | *VPS25* | 0.009(0.005,0.012) | 2.28E-06 | -0.059(-0.19,0.07) | 0.377 | 0.52(-0.01,1.05) | 0.056 | DNAm to T2D |
| cg09185884 | 17 | *KCTD2* | 0.011(0.006,0.015) | 2.33E-06 | 0.001(-0.12,0.12) | 0.989 | 0.15(-2.65,2.95) | 0.919 | Inconclusive - bidirectional |

**Supplementary Table 7. (*Continued*)**

| CpG | Chr | Gene | Meta-EWAS of T2D (N= 3,248) Estimate (95%CI) | P | Forward 2SMR (T2D → DNAm) Estimate (95%CI) | P | Reverse 2SMR (DNAm → T2D) Estimate (95%CI) | P | Likely causal direction |
|---|---|---|---|---|---|---|---|---|---|
| cg11024682 | 17 | *SREBF1* | 0.008(0.005,0.011) | 1.33E-06 | 0.054(-0.07,0.18) | 0.414 | -0.18(-0.61,0.25) | 0.408 | T2D to DNAm |
| cg14284506 | 17 | *Open sea* | -0.005(-0.007,-0.003) | 7.31E-06 | -0.033(-0.17,0.11) | 0.640 | NA | NA | Inconclusive - single direction |
| cg18181703 | 17 | *SOCS3* | -0.01(-0.015,-0.006) | 6.20E-06 | 0.075(-0.04,0.19) | 0.217 | -0.23(-0.72,0.25) | 0.341 | DNAm to T2D |
| cg26766064* | 17 | *MIR657* | -0.007(-0.009,-0.004) | 5.17E-06 | -0.1(-0.22,0.02) | 0.096 | NA | NA | T2D to DNAm |
| cg08570691 | 19 | *RPL13AP5* | -0.008(-0.012,-0.005) | 2.78E-06 | -0.07(-0.21,0.07) | 0.327 | NA | NA | Inconclusive - single direction |
| cg11252555 | 19 | *RPL13AP5* | -0.008(-0.011,-0.004) | 7.44E-06 | -0.054(-0.17,0.06) | 0.370 | -0.28(-0.62,0.06) | 0.110 | DNAm to T2D |
| cg24704287† | 19 | *MIR23A* | -0.011(-0.015,-0.007) | 2.34E-08 | -0.009(-0.13,0.11) | 0.877 | NA | NA | Inconclusive - single direction |
| cg06039489 | 20 | *C20orf26* | 0.016(0.009,0.022) | 2.71E-06 | -0.105(-0.22,0.01) | 0.080 | -0.07(-0.28,0.14) | 0.512 | T2D to DNAm |
| cg14003143 | 20 | *SGK2* | -0.006(-0.008,-0.003) | 4.12E-06 | 0.024(-0.1,0.15) | 0.710 | NA | NA | Inconclusive - single direction |
| cg06500161† | 21 | *ABCG1* | 0.013(0.009,0.017) | 2.34E-11 | -0.067(-0.18,0.05) | 0.263 | -0.16(-0.65,0.32) | 0.512 | Inconclusive - bidirectional |
| cg27037013 | 21 | *Open sea* | -0.015(-0.021,-0.009) | 2.90E-06 | -0.044(-0.16,0.07) | 0.465 | -0.30(-1.04,0.44) | 0.423 | T2D to DNAm |
| cg27115863 | 22 | *Open sea* | -0.011(-0.015,-0.006) | 2.41E-06 | -0.078(-0.2,0.04) | 0.207 | 0.00(-0.15,0.15) | 1.000 | T2D to DNAm |

Meta-EWAS of T2D: meta-analysis of epigenome-wide association studies of prevalent T2D (N=3,428). 2SMR: Two-sample Mendelian randomization. DNAm: DNA methylation. Associations were considered significant at $P < 1.33\times10^{-7}$ in the meta-EWAS of T2D, $P < 0.001$ in the forward 2SMR, and $P < 0.002$ in the reverse 2SMR analysis. * CpGs identified with borderline significance in the forward 2SMR. ** CpGs identified with significance or borderline significance in the reverse 2SMR. †CpGs previously reported with epigenome-wide significance in the meta-EWAS of T2D.‡meta-EWAS CpG analyzed bidirectionally in the 2SMR and observed with consistent direction of association between the causal and the observational analysis. NA: missing data.

**Supplementary Figure 1. Summary plots of the forward two-sample Mendelian randomization at the CpG cg20812370 (*PBX1*), previously detected in a meta-EWAS of prevalent T2D.**

A) Scatter plot depicting the effect of 62 SNPs as individual instruments on T2D (x-axis) and on DNA methylation at cg20812370 (*PBX1*) (y-axis). The fitted regression lines represent the combined effect of these SNPs as a single instrument on DNAm at cg20812370 (*PBX1*). The color of the fitted line refers to the specific MR method used. A negative slope was common to all methods, suggesting that T2D was associated with a decrease in inverse-normal transformed residuals of methylation at cg20812370 (*PBX1*). Highlighted in the plot were the SNPs rs10811660 and rs7903146 that we identified with suggestive heterogeneous effects on DNAm at cg20812370. B) Funnel plot showing the causal effect of each SNP (x-axis) against the inverse of the standard error (SE) of the combined causal effect (y-axis). Some asymmetry in the funnel plot was identified due to the extreme negative effect of SNPs rs319598 and rs1359790 on methylation at cg20812370 (*PBX1*), suggesting horizontal pleiotropy. C) Forest plot illustrating the mean causal effect of each T2D SNP on methylation at cg20812370 (*PBX1*). Mean causal effect is depicted by the black point, with the surrounding line corresponding to the 95% confidence interval (95% CI). At the bottom of the plot, the horizontal red lines show the mean and 95% CI of the meta-analyzed causal effect across SNPs using one of five different MR methods. Effect estimates crossing the vertical dashed line set at “0”, or the line of null associations, indicate no evidence of causality for the single SNP, or the combined causal effect. D) Leave-one-out sensitivity analysis showing the total effect of T2D on methylation at cg20812370 (*PBX1*) after sequentially excluding one SNP at a time from the analysis. This analysis helped to identify how robust was the estimate to the effect of individual SNPs, and if the total causal effect was driven by a single instrument. For this analysis, none of the SNPs was driving alone the total causal estimate at cg20812370 (*PBX1*), and the combined effect using the IVW estimate suggested strong evidence for a negative effect of T2D on methylation at the CpG in *PBX1*.

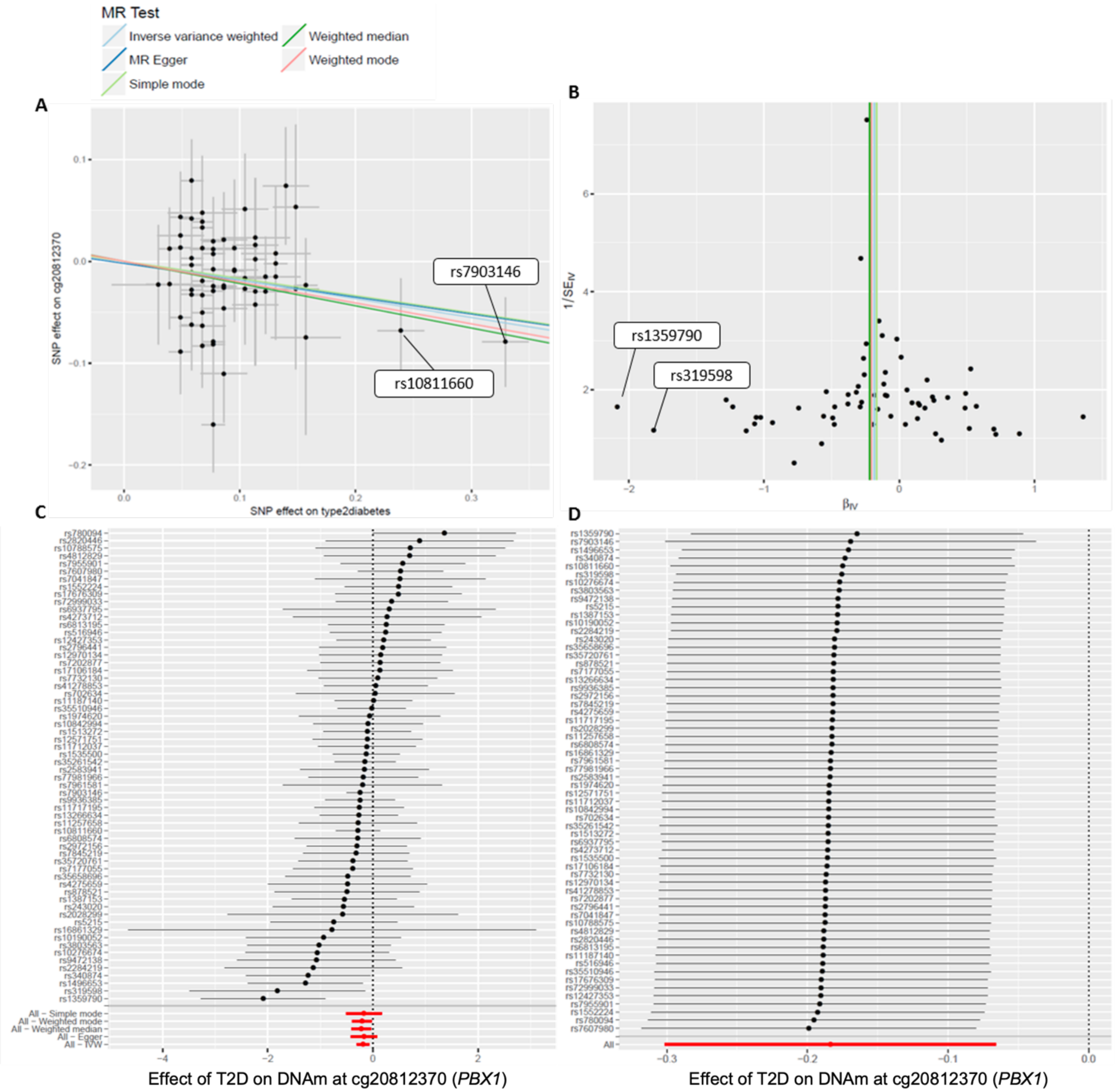
MR Test
Inverse variance weighted
Weighted median
MR Egger
Weighted mode
Simple mode
A
B
C
D
SNP effect on cg20812370
SNP effect on type2diabetes
rs7903146
rs10811660
rs1359790
rs319598
1/SE$_{IV}$
β$_{IV}$
All – Simple mode
All – Weighted mode
All – Weighted median
All – Egger
All – IVW
All
Effect of T2D on DNAm at cg20812370 (*PBX1*)
Effect of T2D on DNAm at cg20812370 (*PBX1*)

**Supplementary Figure 2. Summary plots of the forward two-sample Mendelian randomization at the CpG cg01577083 (*open sea*), previously detected in a meta-EWAS of prevalent T2D.**

A) Scatter plot depicting the effect of 62 SNPs as individual instruments on T2D (x-axis) and on DNA methylation at cg01577083 (*open sea*) (y-axis). The fitted regression lines represent the combined effect of these SNPs as a single instrument on DNAm at cg01577083. The color of the fitted line refers to the specific MR method used. A negative slope was common to all methods, suggesting that T2D was negatively associated with an increase in inverse-normal transformed residuals of methylation at cg01577083. Highlighted in the plot were the SNPs rs10811660 and rs7903146 that we identified with suggestive heterogeneous effects on DNAm at cg01577083. B) Funnel plot showing the causal effect of each SNP (x-axis) against the inverse of the standard error (SE) of the combined causal effect (y-axis). C) Forest plot illustrating the mean causal effect of each T2D SNP on methylation at cg01577083. Mean causal effect is depicted by the black point, with the surrounding line corresponding to the 95% confidence interval (95% CI). At the bottom of the plot, the horizontal red lines show the mean and 95% CI of the meta-analyzed causal effect across SNPs using one of five different MR methods. Effect estimates crossing the vertical dashed line set at "0", or the line of null associations, indicate no evidence of causality for the single SNP, or the combined causal effect. D) Leave-one-out sensitivity analysis showing the total effect of T2D on methylation at cg01577083 after sequentially excluding one SNP at a time from the analysis. For this analysis, none of the SNPs was driving alone the total causal estimate at cg01577083, and the combined effect using the IVW estimate suggested strong evidence for a negative effect of T2D on methylation at this CpG.

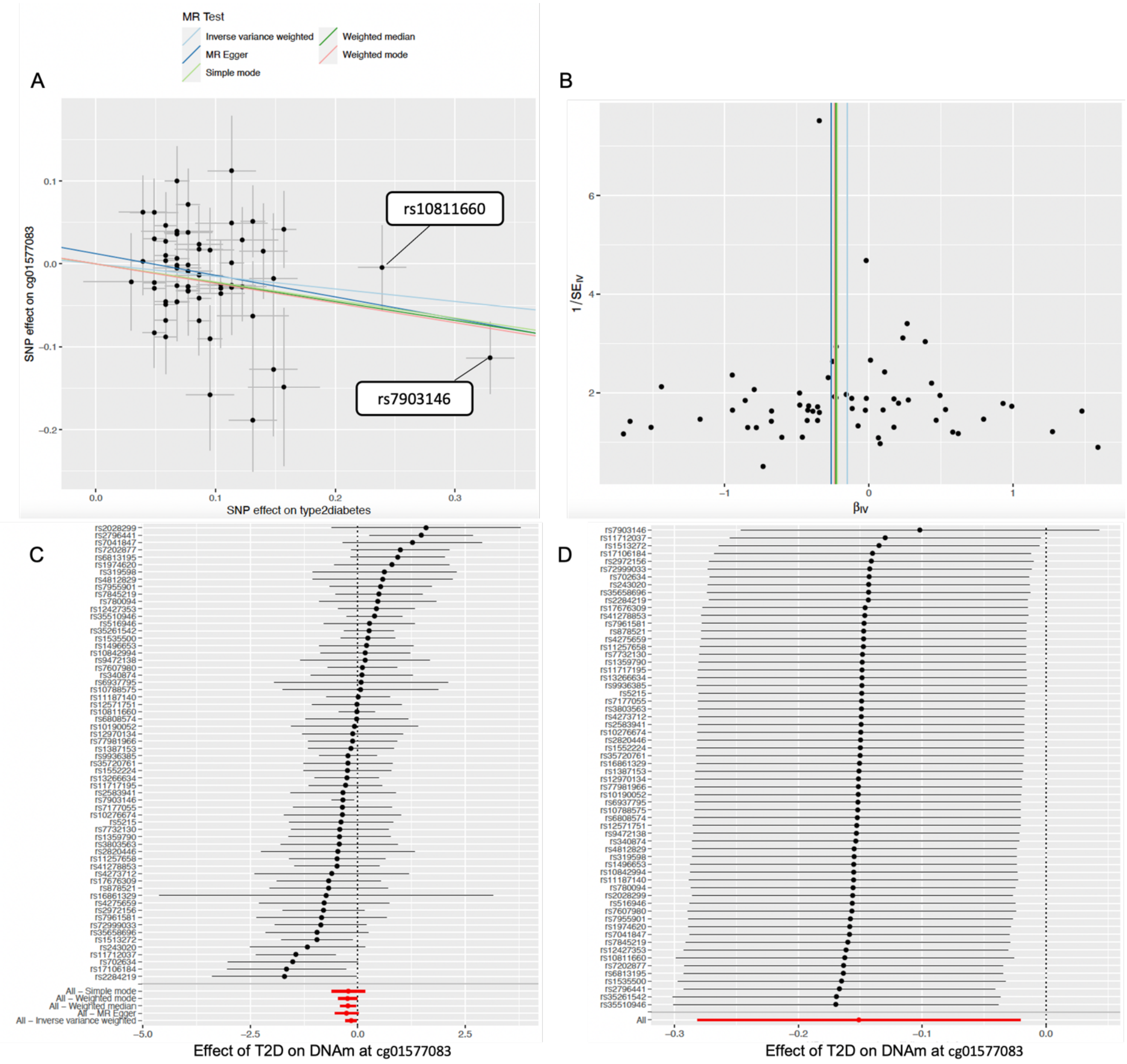

MR Test
Inverse variance weighted
MR Egger
Simple mode
Weighted median
Weighted mode
A
B
rs10811660
rs7903146
SNP effect on cg01577083
SNP effect on type2diabetes
1/SE_IV
β_IV
C
D
Effect of T2D on DNAm at cg01577083
Effect of T2D on DNAm at cg01577083

**Supplementary Figure 3. Forest plot showing enrichment for outcomes listed in the EWAS catalog based on CpGs detected previously in a meta-EWAS of T2D that were analyzed using a bidirectional two-sample MR (2SMR). Results of the enrichment analysis are shown based on the likely direction of association between meta-EWAS CpGs and T2D according to results of the bidirectional 2SMR.**

**CpGs identified as secondary to the effects of T2D (forward 2SMR) (n=12)**

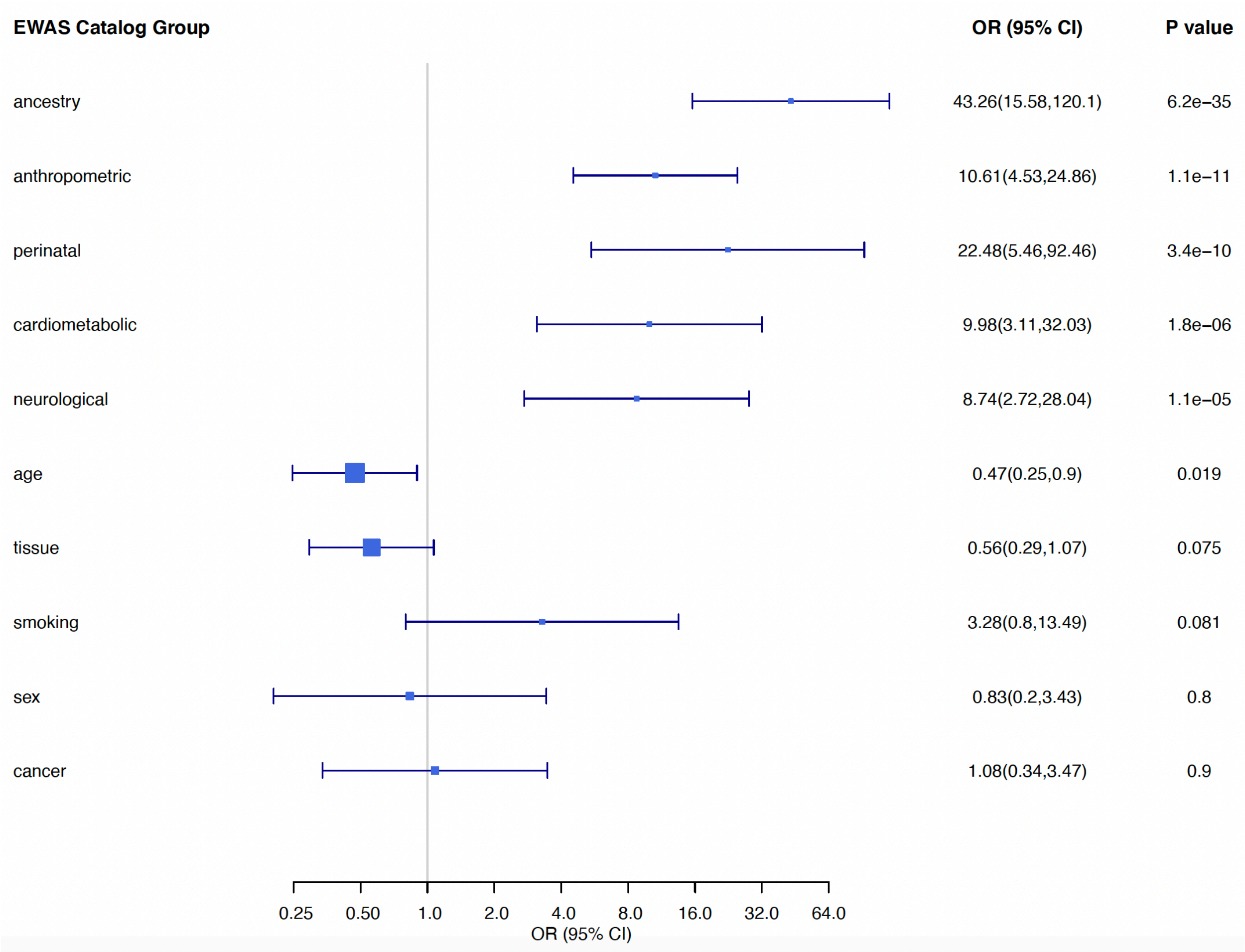

**CpGs identified with a likely causal effect on prevalent T2D (reverse 2SMR) (n=15)**

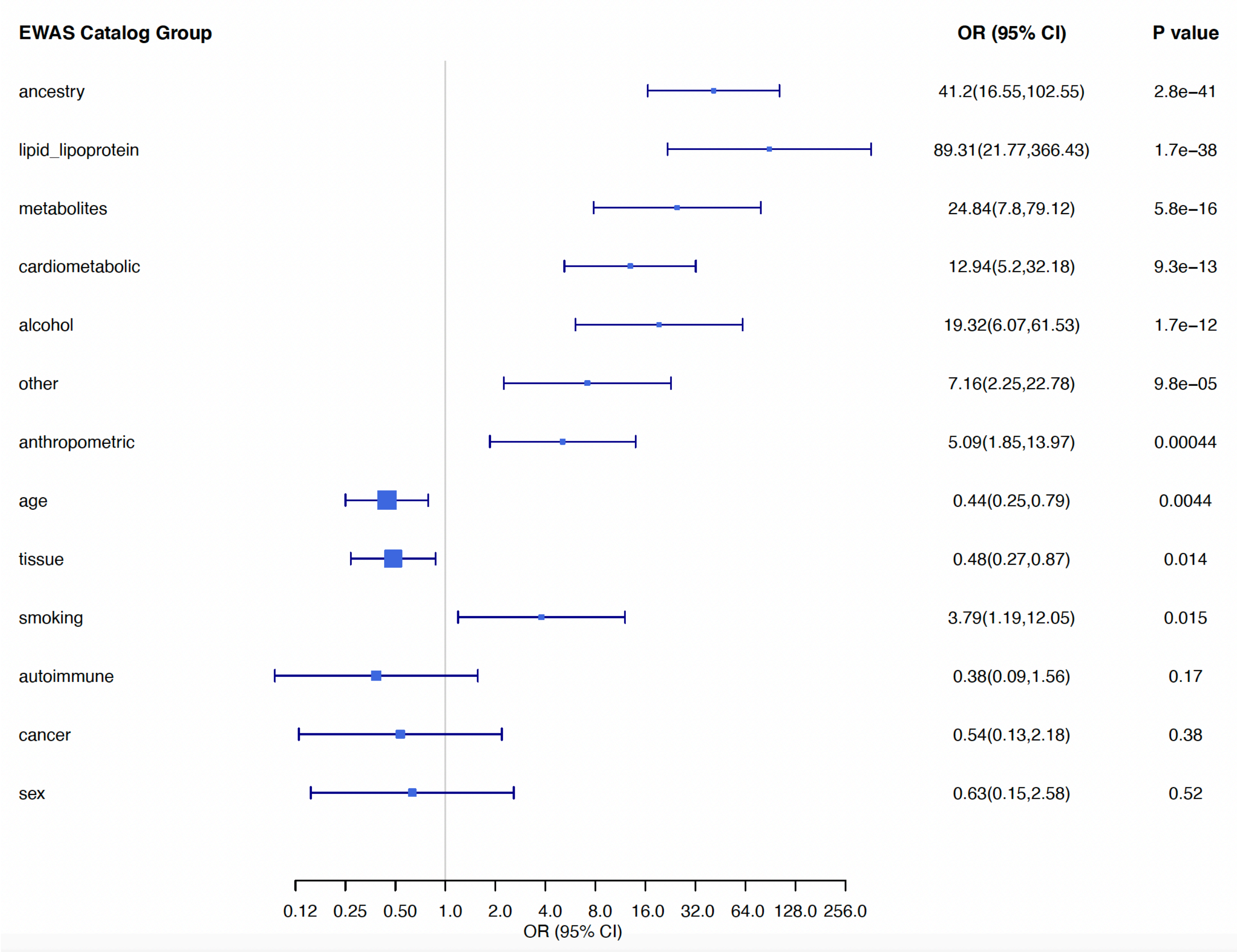

**CpGs with inconclusive direction of association with prevalent T2D based on bidirectional 2SMR data (n=31)**

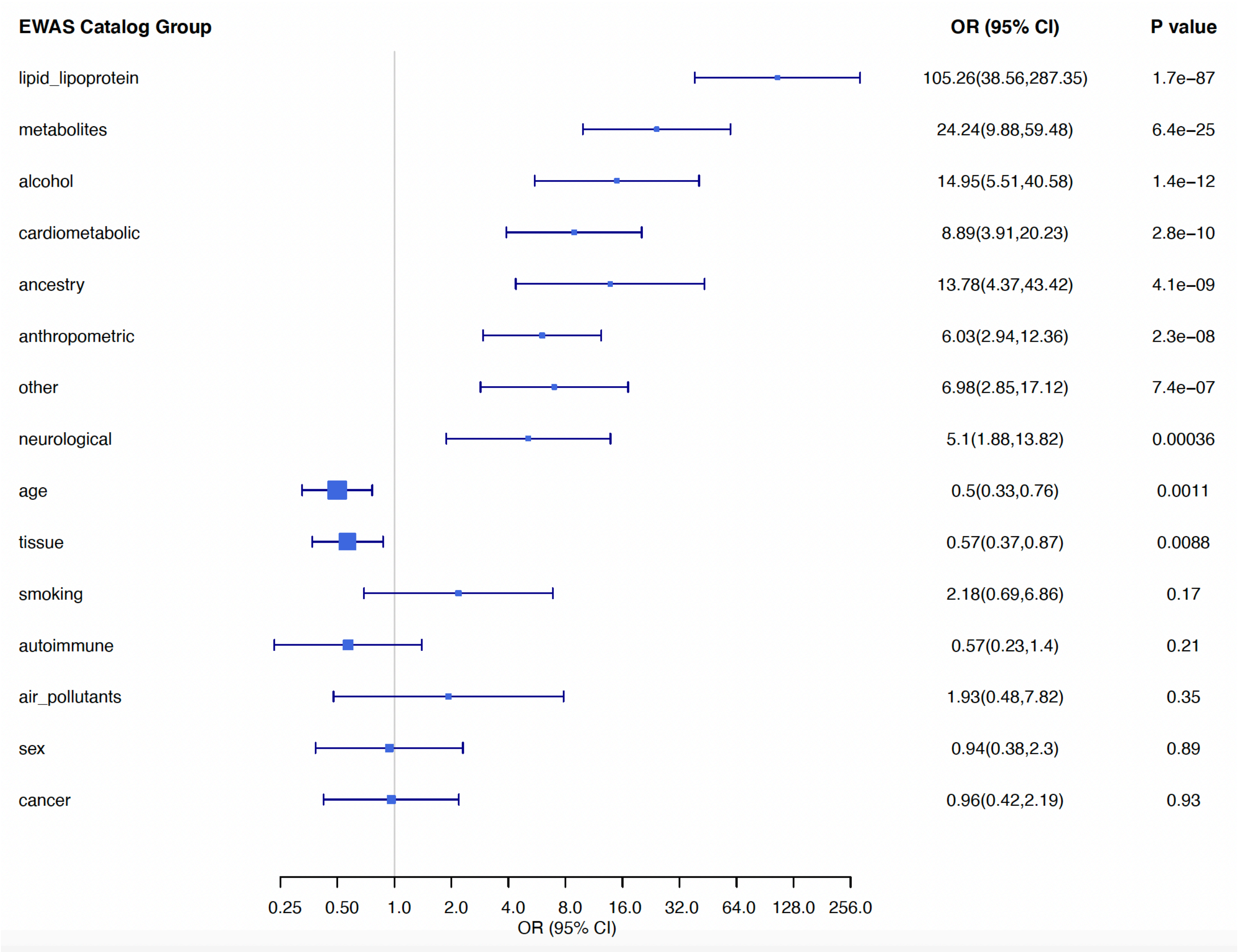


**Ancestry:** ethnicity. **Anthropometric:** bmi, body_mass_index, waist_circumference, arm_circumference, head_circumference, hip_circumference, fat, mass, weight, height, skinfold, waist, bone_mineral_density, head, hip, leg, pelvis, ribs, spine, total_body, trunk, alpha_neck_angle__hip_measurement, theta_neck_angle__hip_measurement, arm_area and arm_bone_mineral_content. **Autoimmune:** rheumatoid, ulcerative_colitis, lupus, crohn, inflammatory_bowel_disease, atopy, graves, psoriasis, multiple_sclerosis, cow_milk_allergy. **Perinatal:** perinatal, birth_weight, birthweight, maternal_underweight, plasma_folate, prenatal, pregnancy, preterm_birth, season_of_birth, breastfeeding, utero, fetal_intolerance_of_labor, Starling-PS_maternal_serum, gestational_weight_gain, parity and fetal_brain_development. **Cardiometabolic:** blood_pressure, diabetes, chronic_kidney_disease, atrial_fibrillation, ischaemic_stroke, hypertension, myocardial_infarction, coronary_heart_disease, obesity, hepatic_fat, statin_use, creactive, c-reactive_protein, insulin, glucose, homair, resistin, hba1c, adiponectin, leptin, liver_fat, proinsulin, arterial_distensibility, common_carotid_intima-media_thickness and pulse_rate. **Lipid lipoproteins:** triglycerides, hdl, highdensity_lipoprotein, lipemia, cholesterol, lipoprotein, ldl, vldl, idl, phospholipids, lp_a, concentration_of_idl_particles, total_lipids_in_idl and total_phosphoglycerides. **Neurological:** dementia, schizophrenia, palsy, alzheimer, depressive_disorder, depressive_symptoms, attention_deficit_hyperactivity_disorder, wellbeing, amyloid_plaques, depression, cognitive, personality_disorder, tic_disorders, aggressive_behaviour, infant_attention, cortical, stress, anxiety, neurobehavioural_scale, seizures, conduct_problems, parkinson, social_communication_deficits, hippocampus_volume, thalamus_volume, antidepressant_use, response_to_antidepressants, apolipoprotein and apoe. **"Other"** correspond to traits not included in age, tissue, smoking, alcohol, sex, ancestry, cancer, autoimmune, infection, cardiometabolic, perinatal, lung, lipid_lipoprotein, anthropometric, neurological, socioeconomic position, social_adversity, diet_environment, air_pollutants, metabolites and miRNA. **Tissue:** tissue.
